# The innate and adaptive immune landscape of SARS-CoV-2-associated Multisystem Inflammatory Syndrome in Children (MIS-C) from acute disease to recovery

**DOI:** 10.1101/2020.08.06.20164848

**Authors:** Eleni Syrimi, Eanna Fennell, Alex Richter, Pavle Vrljicak, Richard Stark, Sascha Ott, Paul G Murray, Eslam Al-Abadi, Ashish Chikermane, Pamela Dawson, Scott Hackett, Deepthi Jyothish, Hari Krishnan Kanthimathinathan, Sean Monaghan, Prasad Nagakumar, Barnaby R Scholefield, Steven Welch, Naeem Khan, Sian Faustini, Pamela Kearns, Graham S Taylor

**Affiliations:** Institute of Immunology and Immunotherapy, University of Birmingham, Birmingham, B15 2TT, UK; Health Research Institute and the Bernal Institute, University of Limerick, Limerick, Ireland; Clinical Immunology Service, Institute of Immunology and Immunotherapy, University of Birmingham, Birmingham, B15 2TT, UK; Warwick Medical School, University of Warwick, Coventry, UK; Bioinformatics Research Technology Platform, University of Warwick, Coventry, UK; Birmingham Women’s and Children’s NHS Foundation Trust, Birmingham, UK; Heartlands Hospital, University Hospitals Birmingham NHS Foundation Trust, Birmingham, UK; Institute of Inflammation and Ageing, University of Birmingham, Birmingham, UK; NIHR Birmingham Biomedical Research Centre and Institute of Cancer and Genomic Sciences, University of Birmingham, Birmingham, UK

## Abstract

Multisystem inflammatory syndrome in children (MIS-C) is a life-threatening disease occurring several weeks after severe acute respiratory syndrome coronavirus 2 (SARS-CoV-2) infection. MIS-C has overlapping clinical features with Kawasaki Disease (KD), a rare childhood vasculitis. MIS-C therapy is largely based on KD treatment protocols but whether these diseases share underpinning immunological perturbations is unknown. We performed deep immune profiling on blood samples from healthy children and patients with MIS-C or KD. Acute MIS-C patients had highly activated neutrophils, classical monocytes and memory CD8+ T-cells; increased frequencies of B-cell plasmablasts and CD27-IgD-double-negative B-cells; and increased levels of pro-inflammatory (IL6, IL18, IP10, MCP1) but also anti-inflammatory (IL-10, IL1-RA, sTNFR1, sTNFR2) cytokines. Increased neutrophil count correlated with inflammation,cardiac dysfunction and disease severity. Two days after intravenous immunoglobulin (IVIG) treatment, MIS-C patients had increased CD163 expression on monocytes, expansion of a novel population of immature neutrophils, and decreased levels of pro- and anti-inflammatory cytokines in the blood accompanied by a transient increase in arginase in some patients. Our data show MIS-C and KD share substantial immunopathology and identify potential new mechanisms of action for IVIG, a widely used anti-inflammatory drug used to treat MIS-C, KD and other inflammatory diseases.

## Introduction

Infection of children with SARS-CoV-2, the viral cause of coronavirus disease 2019 (COVID-19) is associated with two distinct outcomes. The first is an acute infection of the respiratory tract that in most cases is asymptomatic or associated with mild respiratory symptoms^1^. The second is a rare, severe hyperinflammatory syndrome called Multisystem Inflammatory Syndrome in Children (MIS-C) by the World Health Organisation or paediatric inflammatory multisystem inflammatory syndrome temporally associated with SARS-CoV-2 infection (PIMS-TS) in the UK^2–5^. Children with MIS-C present with fever, inflammation and evidence of single or multi-organ failure that manifests with cardiac dysfunction, hypotension and life-threatening shock, several weeks after the primary infection. This is accompanied by lymphopaenia and neutrophilia, both of which are rare in acute paediatric COVID-19^1–4^.

MIS-C does, however, share clinical features with several paediatric inflammatory conditions including toxic shock syndrome (TSS), macrophage activation syndrome (MAS) and Kawasaki Disease (KD). KD is a systemic vasculitis that presents with symptoms of fever, rash, conjunctivitis, lymphadenopathy and cardiac complications and is believed to be triggered by an as yet unidentified infectious agent^6,7^. Acutely ill KD patients have increased blood levels of both pro-inflammatory anti anti-inflammatory cytokines and have lymphopaenia and neutrophilia^7^. Untreated KD can cause coronary aneurysms but the risk is substantially reduced by treatment with intravenous immunoglobulin (IVIG)^7^. This agent is used to treat a diverse range of autoimmune and inflammatory conditions but its mechanism of action remains poorly defined^8,9^.

While MIS-C shares features with KD there are also notable differences^10,11^. For example, MIS-C patients often present with shock, cardiac dysfunction and hyperferritaemia, all of which are rarely seen in KD^2,3,7^. These differences suggest probable differences in the underpinning pathology. The optimal treatment strategy for MIS-C is not known and a range of anti-inflammatory agents have been deployed based on KD and adult COVID protocols^2,3,7,12^. IVIG is widely used, often in combination with steroids, but a range of biological agents targeting the interleukin (IL) IL-6, IL1-β or Tumour Necrosis Factor alpha (TNF-α) pathways have also been used^2,3,10,11,13–16^. Identifying the immunological changes occurring in MIS-C patients, and how these relate to other paediatric inflammatory conditions, is important both to understand the pathogenesis of this new disease and inform rational treatment selection. With this in mind, we undertook a detailed high-dimensional immune characterisation of MIS-C patients presenting at a tertiary paediatric hospital, comparing them to KD patients and healthy children recruited within the same time period. Our results provide new insights into the immunopathology of MIS-C and how immunomodulation resolves the inflammatory process.

## Results

Between late April and October 2020 at Birmingham Women and Children’s Hospital, a tertiary level paediatric hospital, we recruited 16 children meeting the MIS-C diagnostic criteria established by the UK Royal College of Paediatrics and Child Health (**SI Table 1**). A surge of cases (50%) occurred approximately four weeks after SARS-CoV-2 cases were detected in the local community; subsequent cases accrued steadily over time (**Figure 1A**). All 16 MIS-C cases were over 5 years of age and tested positive for anti-SARS-CoV2 antibodies (**Figure 1B and SI Table 1**)^17^. Contemporaneously we recruited two patients with KD both of whom were under 5 years old and negative for anti-SARS-CoV-2 antibodies. Clinical laboratory tests showed all patients had elevated inflammatory markers, including ferritin (**Figure 1C**). Almost all MIS-C patients had high troponin and N-terminal pro B-type natriuretic peptide (NTpro-BNP, data not shown) (**SI-Table 1**). Interestingly, all MIS-C patients were deficient in vitamin D. Patients’ length of hospital stay ranged between 5 to 16 days and 88% of MIS-C patients were admitted to the paediatric intensive care unit (PICU) from 2 to 8 days (**Figure 1D**). Of the 16 MIS-C patients, 13 received IVIG with three MIS-C patients (and one KD patient) receiving a second IVIG infusion due to ongoing inflammation. Eight MIS-C patients received IV methyprednisolone and one, patient 13, received anti-IL6 therapy (Tocilizumab). All 18 patients survived without long term cardiac complications at the time of last follow up.

**Figure 1.**
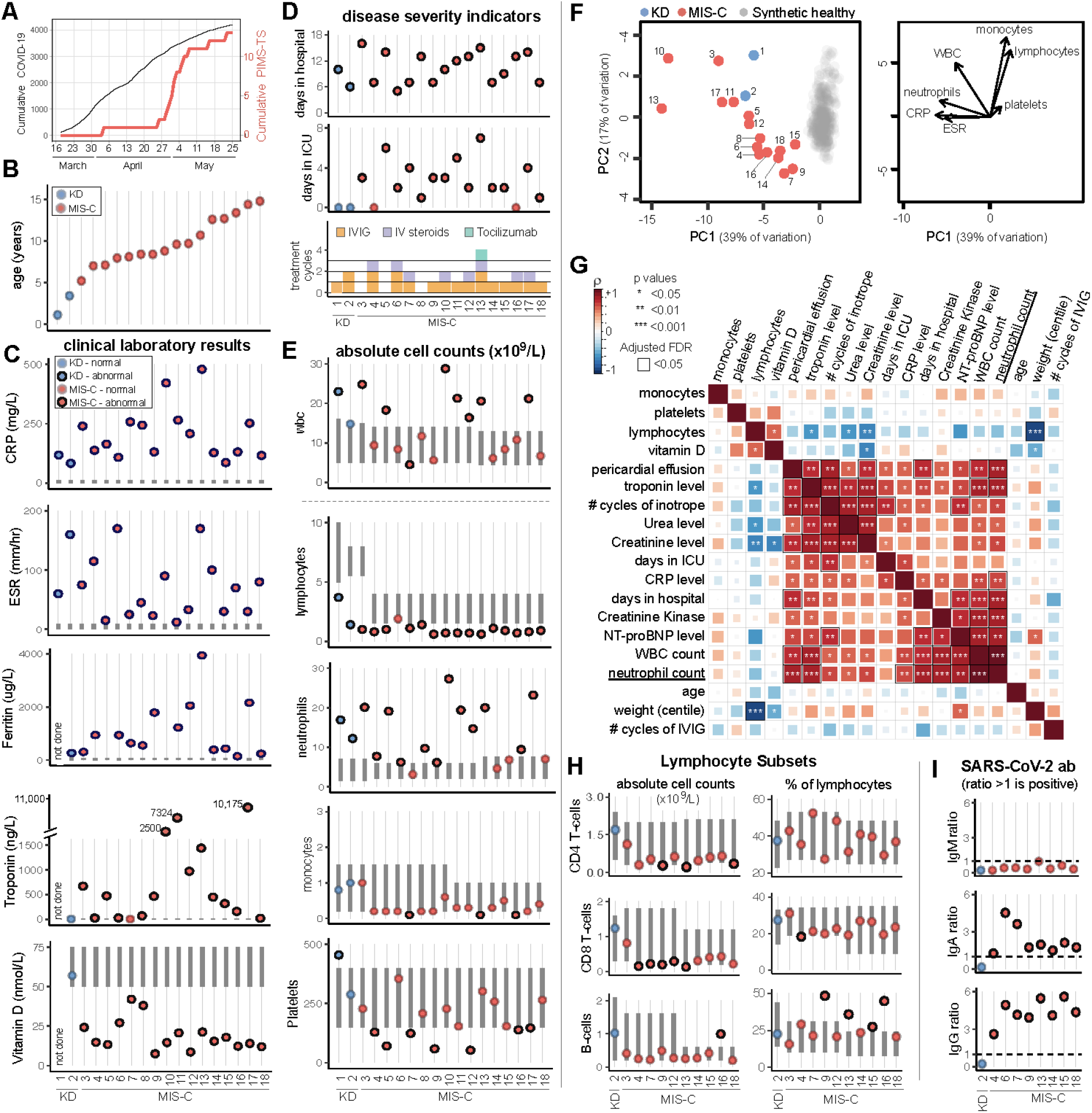
Demographic, clinical and immunological status of 18 paediatric patients with Kawasaki disease or MIS-C. **A**. Cumulative SARS-CoV-2 positive cases identified by PCR testing within the Birmingham area compared to MIS-C cases admitted to Birmingham Children’s Hospital PICU. **B**. Age of KD and MIS-C patients recruited to this study. **C**. Clinical laboratory results shown for C-reactive protein (CRP), erythrocyte sedimentation rate (ESR), ferritin, troponin and vitamin D. **D**. Disease severity indicators shown as days hospitalised, days in PICU and treatment cycles of IVIG, intravenous steroids and Tocilizumab. **E**. Pre-treatment absolute count of different immune cell subsets expressed as 10^9^ cells/L. **F**. Left: Principal component analysis biplot of clinical laboratory features for patients with MIS-C or KD and synthetic healthy controls derived from normal range data. Right: Loading plot showing the top 7 features contributing to principal components one and two. **G**. Correlation matrix of clinical features, immune parameters and demographics for the 16 MIS-C patients. The strength of each correlation is indicated by colour and statistical significance by asterisks: *p<0.05, **P<0.01, ***P<0.001. Black outline indicates a significant result after 5% false discovery rate correction using the Benjamini-Hochberg method. **H**. Pre-treatment frequency of lymphocyte subsets expressed as the absolute number of cells ×10^9^/L (left column) or percentage of total lymphocytes (right column). **I**. SARS-CoV-2 ab responses for IgM, IgA and IgG. In panels C, E and H the normal range for each patient, based on their age, is shown by the vertical dark grey bar. Data points outside the normal range are drawn with a black outline.

### Clinical laboratory data shows neutrophilia is associated with severe disease in MIS-C

In accordance with previous reports^2,3,7,18^, both KD patients and almost all MIS-C patients had abnormally low lymphocyte and abnormally high neutrophils counts (**Figure 1E**). The absolute number of monocytes was normal for the KD patients but at the lower limit of normal for almost all MIS-C patients. Analysis of longitudinal data showed these perturbations resolved following treatment, although this was slower for granulocytes which were still elevated once lymphocytes and monocytes had returned to normal (**SI Figure 1**). Using principal component analysis to obtain a global view of the clinical laboratory data showed neutrophil count, CRP and ESR correlated with each other with monocyte and lymphocyte counts orthogonal to these features (**Figure 1F**). All patients fell outside of the normal region to varying degrees.

We next investigated the relationships between clinical features, absolute immune cell counts and demographics for the MIS-C patients (**Figure 1G**)^19^. As expected, clinical markers of cardiac and kidney dysfunction (troponin, pericardial effusion, urea and creatinine) positively correlated with the need for inotrope support. We observed multiple highly significant positive correlations between absolute neutrophil count and markers of inflammation (CRP), cardiac dysfunction (presence of pericardial effusion, levels of troponin, creatinine kinase and NTpro-BNP) and overall length of hospital stay. Additional clinical laboratory assays performed on a subset of patients showed that, in accordance with lymphopaenia, the absolute counts of CD4 and CD8 T-cells and B-cells were diminished. However, the relative proportion of these cells within the lymphocyte pool were generally unaltered (**Figure 1H**). Analysing the anti-SARS-CoV-2 antibody response in detail for eight MIS-C patients showed all eight had IgG and IgA antibodies but lacked IgM antibodies consistent with MIS-C developing weeks after virus infection occurred (**Figure 1I**)^1,2,3^.

### scRNA sequencing data show monocytes are profoundly altered in MIS-C and KD patients

To explore immune changes in MIS-C in more detail, we first performed an unbiased analysis by performing single cell RNA sequencing (scRNAseq) on the pre-treatment peripheral blood mononuclear cells (PMBCs) of two representative patients (P13 and P14) with MIS-C. Both patients were admitted to PICU but patient P14 responded rapidly to one cycle of IVIG and stayed on PICU for three days whereas patient P13 stayed on PICU for eight days and received a second IVIG infusion, intravenous steroids and Tocilizumab, a monoclonal antibody against the interleukin-6 receptor, to control their disease. A convalescent sample from P13 collected at discharge from PICU was also analysed. For comparative purposes we also analysed pre-treatment acute-stage PBMCs from patient KD2 who required two infusions of IVIG (**Figure 1D**). Unsupervised clustering of 10,031 cells produced 19 different clusters comprising all major lymphocyte subsets (**Figure 2A, SI Fig 2 and 3**). Analysing each patient separately, we observed that all possessed lymphocytes assigned to one of the nine B-cell, T-cell or NK cell clusters, although the frequencies varied between patients and for P13 from acute disease to convalescence (**Figure 2B and 2C**). In contrast, each patient’s monocytes were assigned to only one or two of the five different monocyte clusters. Four of these clusters corresponded to CD14+ classical monocytes and for each sample these cells were assigned to a separate cluster indicating there were considerable inter-sample differences in gene expression. Based on CD14 and CD16 expression the fifth monocyte cluster (cluster nine) contained intermediate and non-classical monocytes, and these varied markedly in frequency between samples: abundant in the acute sample from patient KD2, less frequent in the acute sample from patient P13 and scarce in the convalescent sample from this same patient but also the acute sample from P14 Pathway analysis (**Figure 2D**) on genes upregulated in each of the major immune subsets, relative to the same subset in the convalescent sample from P13, showed highly significant enrichment of genes annotated with GO-term 0002446 ‘Neutrophil mediated immunity’ in CD14+ monocytes from patient KD2 (p=3×10^−34^) and P13 (p=7×10^−31^). Although labelled as neutrophil mediated immunity, this pathway contains many genes expressed by monocytes. Genes upregulated in CD14+ monocytes from KD2 and P13 included: genes involved in complement function and regulation (CD35 and CD55); adhesion, homing and scavenger receptors (CD36, CD62L, CD63); Fc receptors (FCGR2A and FCER1G); alarmin related S100A molecules (S100A8, S100A9, S100A11 and S100A12) and regulation of innate cell mediated inflammation (SERPINB1). Due to the lack of intermediate/non-classical monocytes in the convalescent sample we did not perform GO-term analysis for this subset.

**Figure 2.**
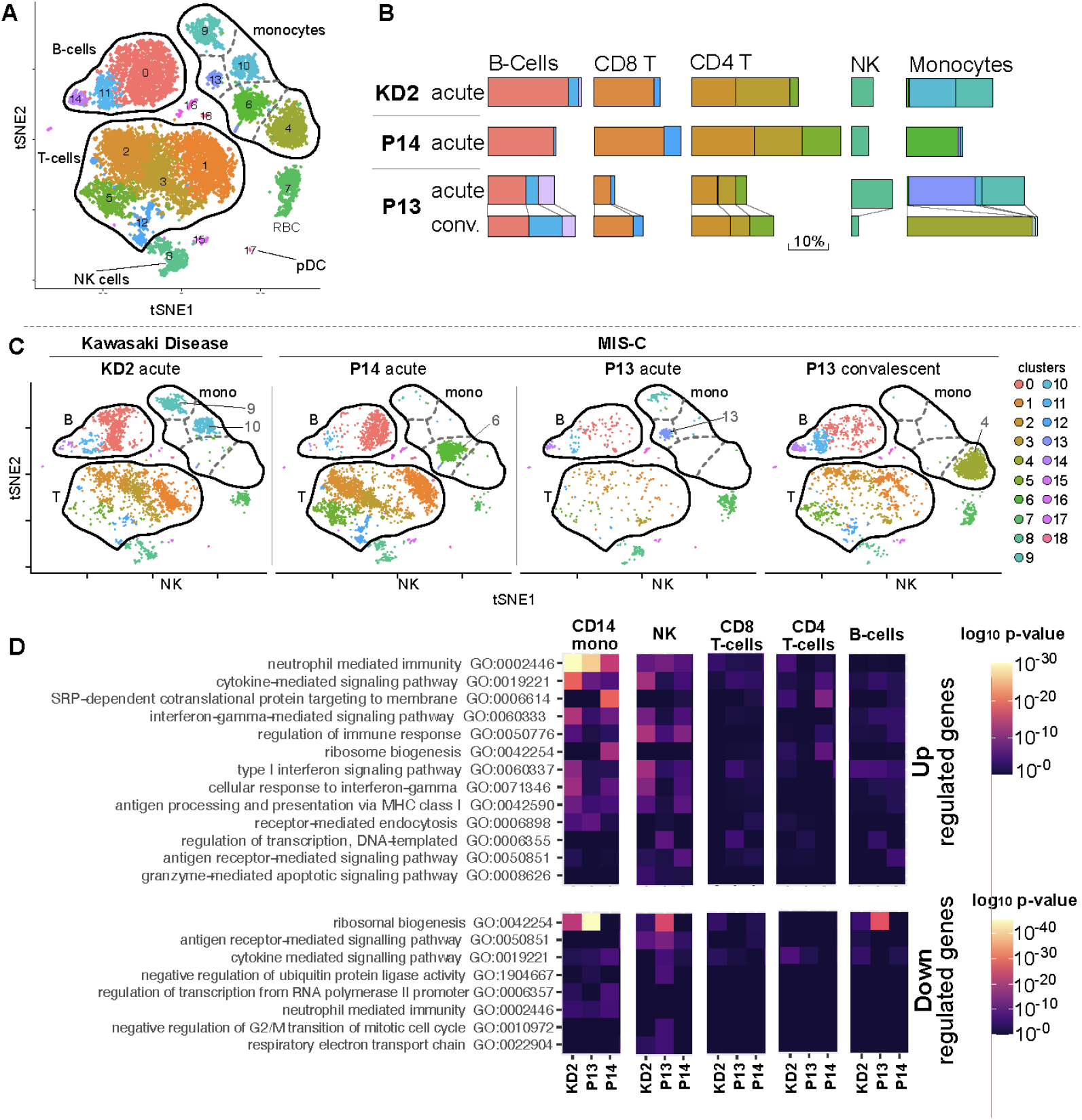
Single cell RNA sequence analysis of MIS-C and KD PBMC. **A**. tSNE representation of major cell types and associated FlowSOM clusters in acute stage PBMCs from patients KD2, P13, P14 and a convalescent sample from P13 at discharge from PICU. **B**. Relative size of PBMC clusters for each patient sample. **C**. tSNE representation of PBMCs from each patient sample. **D**. Gene set enrichment analysis of genes upregulated (top panel) or downregulated (bottom panel) for each of the indicated immune cell subsets in acute samples from patients KD2, P13, P14 relative to the convalescent sample from P13. The significance of each pathway is indicated by colour.

The marked changes we observed in monocyte cluster assignment and frequency between the four different samples led us to explore this subset in more detail. Repeating dimensionality reduction and clustering analyses on just the subset of data from the four samples that represented monocytes yielded nine monocyte clusters (numbered mc0 to mc8, **Figure 3A**) that could now be assigned to the three canonical monocyte subsets (**SI Figure 5**). Clusters mc0, mc1, mc3, mc4 and mc5 were CD14^+^ CD16^-^ classical monocytes (CM), mc6 (CD14^int^ CD16^int^) comprised intermediate monocytes (IM) while mc2 (CD14^lo^ CD16^hi^) comprised non-classical monocytes (NCM). Previous studies have shown that the large majority of monocytes in the blood of healthy children are CM with only small populations of IM and NCM present^20^.This distribution was distorted in the acute KD sample with high frequencies of IM (cluster mc6, 8%) and NCM (cluster mc2, 38%) within the total monocyte population (**Figure 3B**). The acute sample from MIS-C patient P13 also had an abnormally high frequency of NCM (34% of total monocytes) but lacked IM. Upon recovery, the MIS-C patient’s monocytes returned to the normal state with only the CM population present in their convalescent sample. Examining gene expression in more detail (**Figure 3C and SI Fig 6**) detected mRNA encoding IL1-beta in the mc6 intermediate monocytes abundant in patient KD2 but detected low or no transcripts encoding other monocyte-associated cytokines including IL6, IL8 (gene CXCL8), IL10, IL18, TNF-a or IL1 receptor antagonist (IL1RA, gene name IL1RN).

**Figure 3.**
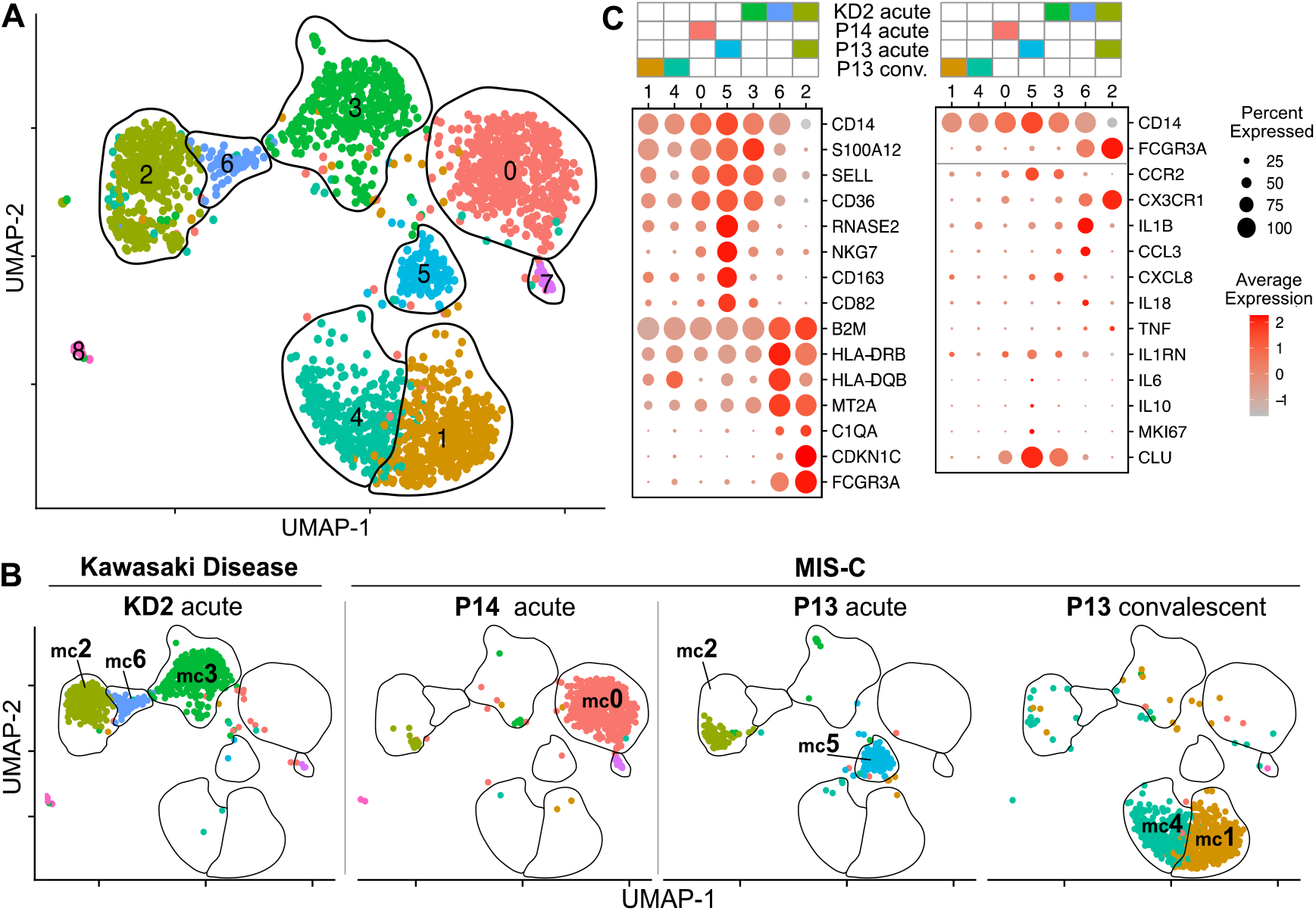
Detailed single cell RNAseq analysis of monocytes. **A**. UMAP representation of monocyte cells after re-clustering on monocytes alone in acute stage samples from patients KD2, P13, P14 and a convalescent sample from P13 at discharge from PICU. **B**. UMAP representation of monocyte clusters from each patient sample. **C**. Expression of select genes in monocyte clusters.

### Mass cytometry shows monocytes granulocytes and CD8 memory T-cells are contemporaneously activated in MIS-C and KD patients

To examine the above changes in more patients and to extend our analysis to granulocytes which we and others have shown are abnormally expanded in MIS-C and KD patients’ blood^2,3^, we developed a 38 marker mass cytometry panel (**SI Table 2**) and used this to investigate whole blood samples from 7 patients (6 MIS-C and 1 KD) and 7 healthy children. The former included patients P13, P14 and KD2 (whose PBMCs were examined by scRNA sequencing) and four additional MIS-C patients from our cohort. Samples were collected during the acute stage, two days after IVIG administration and upon discharge from PICU or hospital **(SI Figure 7**). Acute stage samples were taken before IVIG with one exception, the sample from P14, which was collected 8 hours after IVIG infusion.

Unsupervised dimensionality reduction and clustering of 224,000 cells (16,000 from each of the seven healthy children, six MIS-C patients and one KD patient) identified 24 clusters comprising plasmacytoid dendritic cells (pDCs), T-cells, B-cells, NK-cells and monocytes (**Figure 4A and SI Figure 8**). Comparing the combined data from 7 healthy children to 6 acutely ill patients we observed profound changes in monocyte cluster abundance as noted in the scRNAseq data analysis. We next evaluated the data for each individual (Figure **4B**). Seven clusters were significantly different between MIS-C patients and healthy children. Note that we chose not to include the KD patient in this statistical analysis but show their results on the plot as an exemplar of this disease. Based on marker expression (**SI Figure 8**) the frequency of activated CM (cluster 20) was significantly increased in MIS-C with a concomitant decrease of non-activated CM (cluster 17). There was no significant difference in the frequency of both IM and NCM (cluster 12), but we noted patient P13 and KD2 had high frequencies of these cells as observed in the scRNAseq data. Examining other immune cell types, MIS-C patients showed a small but significant increase in CD19^+^ CD38^hi^ CD27^hi^ B-cell plasmablasts (cluster 5) and a much larger increase in IgD-CD27-double negative (DN) B-cells (cluster 2). These DN B-cells lacked CD11c consistent with them being the recently proposed DN1 B-cell subset^21^. T-cell (cluster 8) and pDC (cluster 11) were both decreased in MIS-C patients relative to control donors.

**Figure 4.**
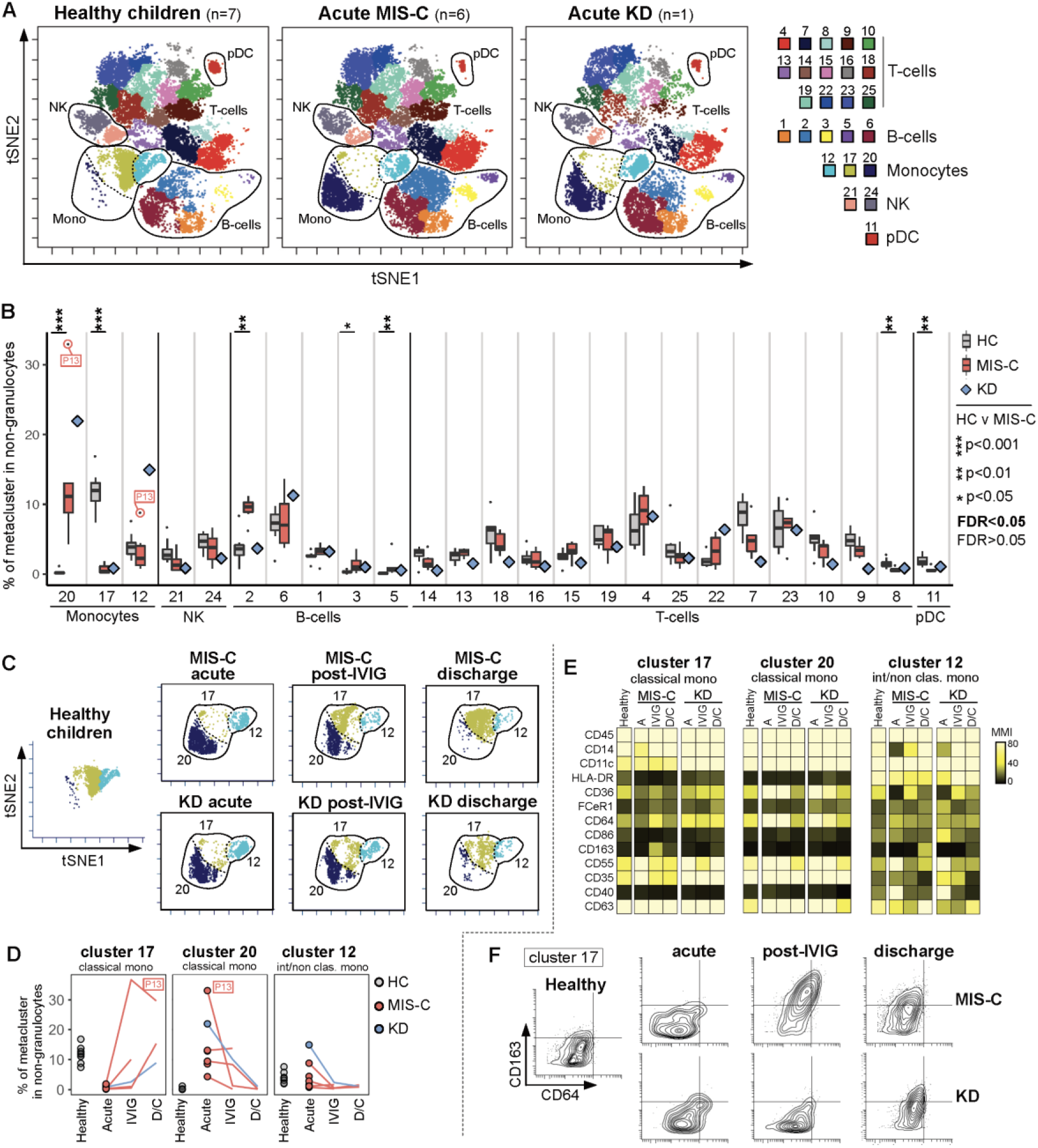
Mass cytometry analysis of mononuclear cells in whole blood samples from healthy children and patients with MIS-C or KD. **A**. tSNE plots of concatenated flow cytometry data from six MIS-C patients or one KD patient at the acute stage of their disease alongside seven healthy children (HC). Each meta-cluster is represented by a different colour and key populations are indicated on the plots. Results from these concatenated datafiles are shown throughout this figure. **B**. The frequency of each FlowSOM metacluster in the same donors expressed as a percentage of total non-granulocyte mononuclear cells are shown as box and whisker plots (healthy volunteer children and MIS-C) or a blue diamond (KD). Results of Wilcoxon rank-sum tests comparing the frequency of each cluster in healthy children and MIS-C patients are indicated by: * p<0.05, ** p<0.01, *** p<0.001. Non-significant results are not shown and emboldened p value symbols indicate significant results after 5% false discovery rate correction using the Benjamini-Hochberg method. **C**. tSNE plots showing cells within monocyte clusters 17, 20 and 12 for MIS-C patients at the acute stage (n=6), post IVIG (n=4) and at discharge (n=2) from PICU alongside a single KD patient and healthy children (n=7). **D**. Trajectory of each of the three monocyte clusters over time in seven healthy children or patients over time (acute stage, post IVIG and PICU discharge). Data from patient P13 is indicated on the plots. **E**. Heatmaps showing the median metal intensity (MMI) of markers expressed on monocyte clusters 17, 20 and 12.**F**. Biaxial plots of CD64 and CD163 expression on cluster 17 monocytes cells in healthy children or patients with MIS-C or KD at the acute, post-IVIG or PICU discharge stages of disease.

We next analysed the post-IVIG and discharge samples available from these patients (SI Figure 7). This showed that CM had started to normalise in the post-IVIG samples, with decreases in the frequency of cells in activated CM and IM/NCM (cluster 20 and 12 respectively) and increases in the frequency of non-activated CM (cluster 17). This reversion to normality continued further to PICU discharge, at which point the patients’ classical monocyte cluster distributions resembled healthy children (**Figure 4C and 4D**). Reversion proceeded rapidly for patient P13, who had the highest frequency of activated CM (cluster 20) at the acute stage. Examining the phenotype of each monocyte cluster over time, we saw increased CD64 and CD163 co-expression on non-activated CM (cluster 17) after IVIG (**Figure 4E**). These dual positive cells were present in 3 of the 4 MIS-C patients who received IVIG and from whom we obtained post-IVIG samples (**SI figure 9**). They were also present in the discharge sample from KD patient KD2, which was collected two days after a second cycle of IVIG was administered (**SI Figure 7**).

Examining the T-cell marker expression data (**SI Figure 8**) we noted most T-cell clusters were CD45RA^+^ CD27^+^ naïve cells. One cluster of CD8 non-naïve T-cells (cluster 13, with low CD45RA and CD27 expression) expressed HLA-DR, a marker of T-cell activation^22^.To explore this in more detail we used CD27 and CD45RA to manually gate CD8+ and CD4+ T-cells into the four canonical T-cell subgroups: naïve (Tn), central memory (Tcm), effector memory (Tem) and terminally differentiated effector memory re-expressing CD45RA (TemRA)^23^ (**SI Figure10)**. As expected for children, and consistent with the unsupervised clustering, we found all donors had a high proportion of naïve T-cells (**Figure 5A**) with the highest proportion in the KD patient likely due to their younger age^24,25^. Comparing healthy children to MIS-C patients we found no significant differences in the distribution of the four subgroups in CD8 or CD4 T-cells (**SI Figure 10C**). However, MIS-C patients had a significantly higher proportion of HLA-DR positive activated CD8-T-cells at the acute stage of disease (**Figure 5B**). In MIS-C patients, only 2% of Tn cells were HLA-DR positive whereas 35% of Tcm and 30% of Tem CD8+ T-cells were HLA-DR positive. The proportion of HLA-DR positive CD8 T-cells remained high after IVIG and, for one patient (P7), increased markedly (**Figure 5C**). At discharge the proportion of activated cells had declined but were still higher than controls. In contrast, only a small proportion of CD4 T-cells expressed HLA-DR and only Tcm were significantly higher than controls at the acute stage. The KD patient showed the same pattern of HLA-DR expression on their CD8+ and CD4+ T-cell subsets.

**Figure 5.**
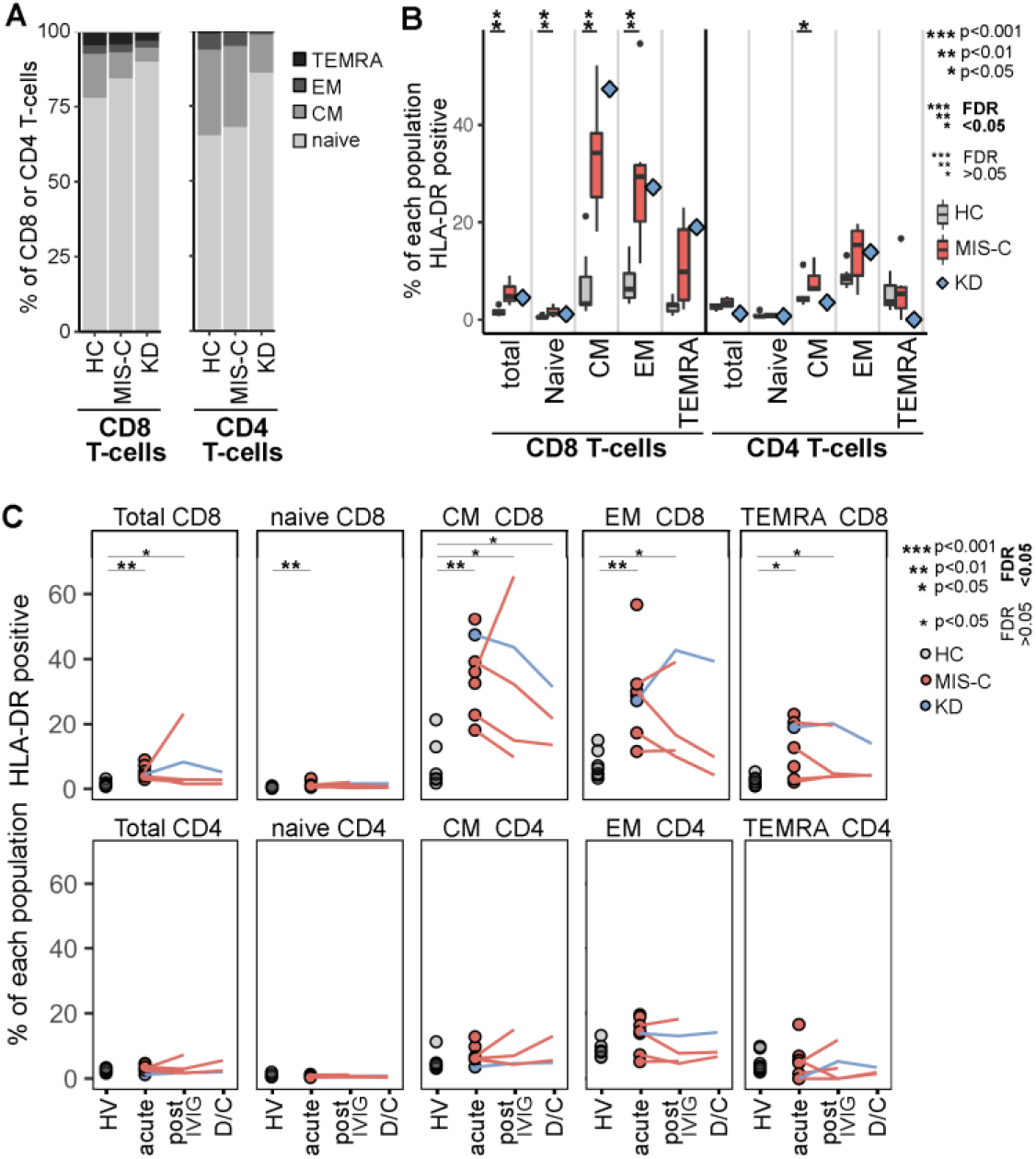
mass cytometry of mononuclear cells analysed by manual gating on canonical T-cell sub-populations. **A**. Percentage of CD8+ and CD4+ T-cells in each of the four canonical T-cell sub-populations for healthy children, and patients at the acute stage of their disease. **B**. Percentage of each T-cell subpopulation from the same donors that were positive for HLA-DR. The results of Wilcoxon ranked sum tests comparing the frequency of each cluster in healthy children to MIS-C patients are shown. **C**. Percentage of each T-cell sub-population positive for HLA-DR over the course of disease. The results of Wilcoxon ranked sum tests comparing the frequency of each cluster in healthy children to MIS-C patients are shown. For all panels the data were from seven healthy children, a single KD patient or MIS-C patients (acute n=6, post-IV n=4, PICU discharge n=2). Significant results are indicated: * p<0.05, ** p<0.01, *** p<0.001. Non-significant results are not shown. Emboldened p value symbols indicate significant results after 5% false discovery rate correction using the Benjamini-Hochberg method.

Turning to granulocytes, we manually gated and examined the CD66b+ CD16+ neutrophil population (**Figure 6A**). We compared equal numbers of cells (22,000) sampled from seven healthy children or from MIS-C patients at each of three key timepoints (6 acutely ill, 4 after IVIG and 2 at time of discharge); for comparison we also examined cells from these same timepoints from KD patient KD2 (**Figure 6B**). Both patient groups exhibited the same changes in phenotype, with decreased expression of the granulocyte maturity markers CD16 and CD10 and increased expression of the neutrophil activation marker CD64^8^, which was highest at the acute stage then slowly decreased after IVIG and at discharge, although levels were still raised at this time^26,8^.

**Figure 6.**
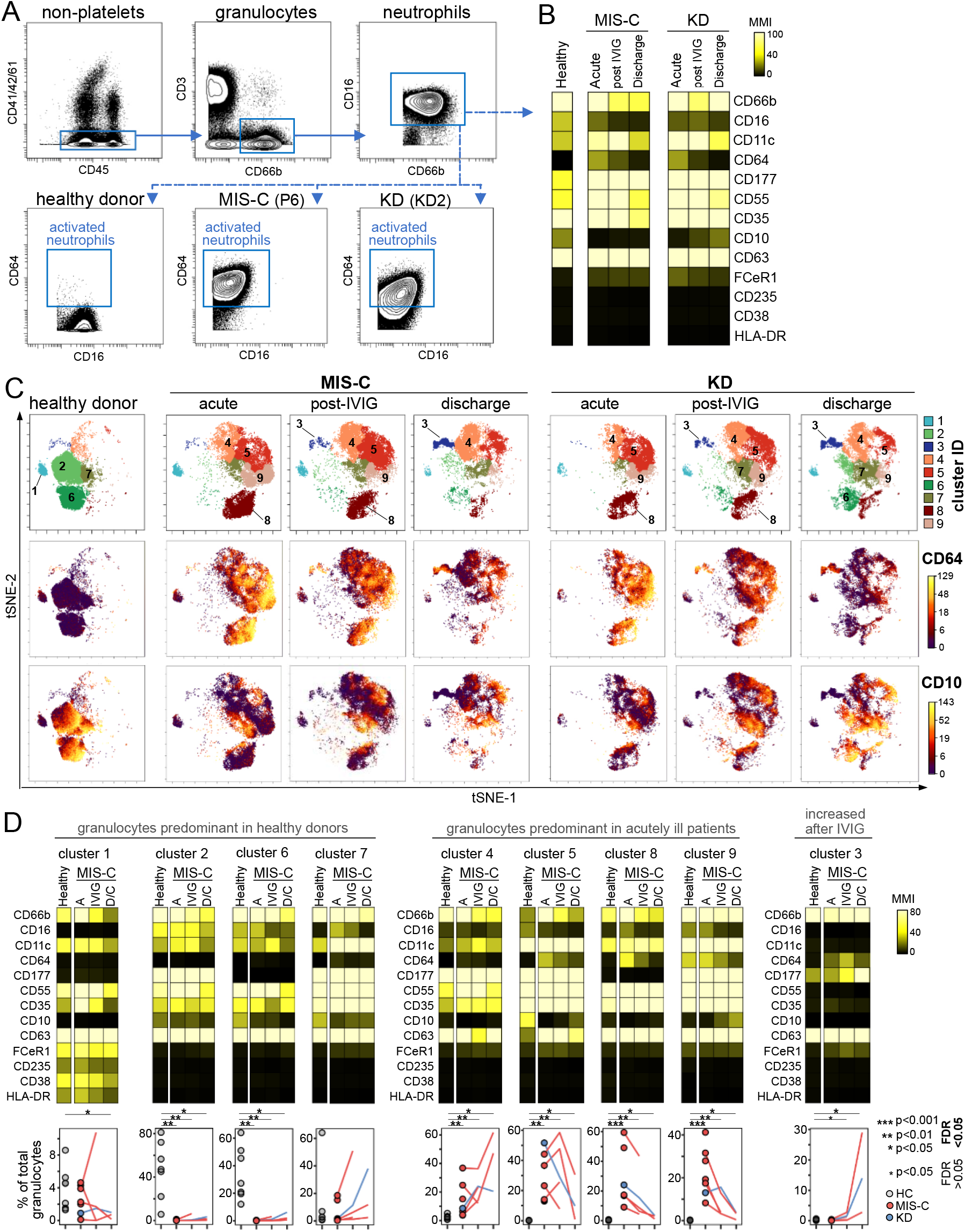
Mass cytometry analysis of granulocytes in whole blood samples from healthy children and patients with MIS-C or KD. **A**. Gating strategy used to manually gate and analyse neutrophil activation in whole blood. **B**. Heatmaps showing median metal intensity (MMI) of markers expressed on manually gated neutrophils. Data are from concatenated FCS files from seven healthy children, MIS-C patients (acute n=6, post-IV n=4, PICU discharge n=2) and a KD patient. **C**. tSNE plots of granulocytes from the same donors analysed by unsupervised clustering. Top row: FlowSom metaclusters. Middle row: CD64 expression. Bottom row: CD10 expression. **D**. Upper panels: heatmaps showing expression level of different markers in each metacluster for the same donors. Lower panels: Trajectory of each metacluster over time, expressed as a percentage of total granulocytes, for each of the healthy children and patients. The results of Wilcoxon ranked sum tests comparing the frequency of each cluster in healthy children to all patients (six MIS-C and one KD patient) at the acute, post-IVIG and PICU discharge timepoints as indicated: * p<0.05, ** p<0.01, *** p<0.001. Non-significant results are not shown and emboldened p value symbols indicate significant results after 5% false discovery rate correction using the Benjamini-Hochberg method.

Unsupervised dimensionality reduction and clustering of the total granulocyte population identified nine clusters (**Figure 6C and 6D**). One cluster, cluster 1, present in patients and healthy children were eosinophils based on expression of FceR1, CD38, HLA-DR and lack of CD16 and CD10 (**Figure 6D**). The other 8 clusters were neutrophils and these showed marked differences in abundance. Healthy children had few cells classified into clusters 4,5,8 and 9 whereas almost all cells in acutely ill MIS-C patients belonged to these clusters. This redistribution of neutrophils was driven by a dramatic decrease in CD10 and increase in CD64 on patients’ neutrophils. Expression of both markers had begun to decrease towards healthy children’s levels after IVIG administration then further decreased at discharge, although the frequencies of all four activated clusters (clusters 4, 5, 8, 9) were still significantly higher at this time. In contrast eosinophils (cluster 1) showed only modest changes in frequency and phenotype with only CD35 (complement receptor 1) expression varying over time.

Granulocyte cluster 3 was present at very low frequency in healthy children (median frequency 0.16% of granulocytes), and in acutely ill MIS-C (0.11%) and KD (0.04%) patients. However, after IVIG the frequency of these cells increased 5 to 12-fold over pre-treatment values in the MIS-C patients (median frequency 0.81% range 0.30-2.7%) and 30-fold (frequency 1.6%) in the KD patient. These cells continued to increase in frequency over time and at discharge their frequency was 70-fold to 204-fold higher than at the acute stage. At discharge they comprised 2.79% and 29.05% of total granulocytes in MIS-C patients P6 and P13 respectively. This continued increase also occurred in the KD patient (KD2) with the frequency of cluster 3 cells 280-fold higher in the discharge sample compared to the acute sample, comprising 14.0% of this patient’s granulocytes at discharge. Cluster 3 cells also possessed an unusual phenotype; they were clearly granulocytes based on their strong expression of the canonical granulocyte marker CD66b with their lack of CD16 and CD10 indicating immaturity. However, they were different to all other granulocyte clusters as they lacked expression of CD11c, CD35 and CD55. Cluster 3 granulocytes in MIS-C patients and the KD patient but not healthy children, expressed CD64 indicating these cells were not only increased in frequency but were also activated over the course of disease.

### Pro- and anti-inflammatory cytokines are elevated in MIS-C and KD patients and arginase is increased after IVIG administration

Finally, we performed a wide-ranging analysis of 32 cytokines and chemokines in plasma samples from nine patients (eight MIS-C, one KD) and seven healthy children. These soluble immune mediators were selected based on the cellular changes we observed in MIS-C and KD, as well as prior studies on KD and, more recently, MIS-C patients^10,11,13–16,27–32^. We selected assays capable of providing absolute quantification to allow data from our patients to be directly compared with historical data from KD and other inflammatory conditions. Comparing MIS-C patients to healthy children, we identified statistically significant differences for 16 of the 32 soluble mediators analysed (**Figure 7A**). MIS-C patients had significantly increased levels of the chemokines monocyte chemoattractant protein 1 (MCP-1/CCL2) and interferon gamma-induced protein 10 (IP10/CXCL10), higher levels of the pro-inflammatory cytokines IL-6 and IL-18 but, at the same time, higher levels of anti-inflammatory cytokine IL-10. Soluble receptors of tumour necrosis factor alpha (sTNF-R1 and sTNFR2), CD40 ligand (sCD40L) and IL-2 (sCD25) were all higher in MIS-C patients as was Interleukin-1 receptor antagonist (IL-1RA), a member of the IL1 family that binds the IL1-receptor to inhibit this pathway. Plasminogen activator inhibitor 1 (PAI-1), pentraxin-3 (PTX3), myeloperoxidase (MPO) and IL-18 were also higher in MIS-C patients. For several pro-inflammatory cytokines and chemokines there was no difference between patients or the healthy donor controls, including: IL1-β, IL-8 (CXCL8), IL-17A, interferon-alpha2 (IFN-a2) interferon-gamma (IFN-ϒ) and TNF-α. We did not include the KD patient in the statistical analysis, but their acute blood sample had the same profile of cytokines, chemokines and other soluble factors as the acute MIS-C patients (**Figure 7A)**.

**Figure 7.**
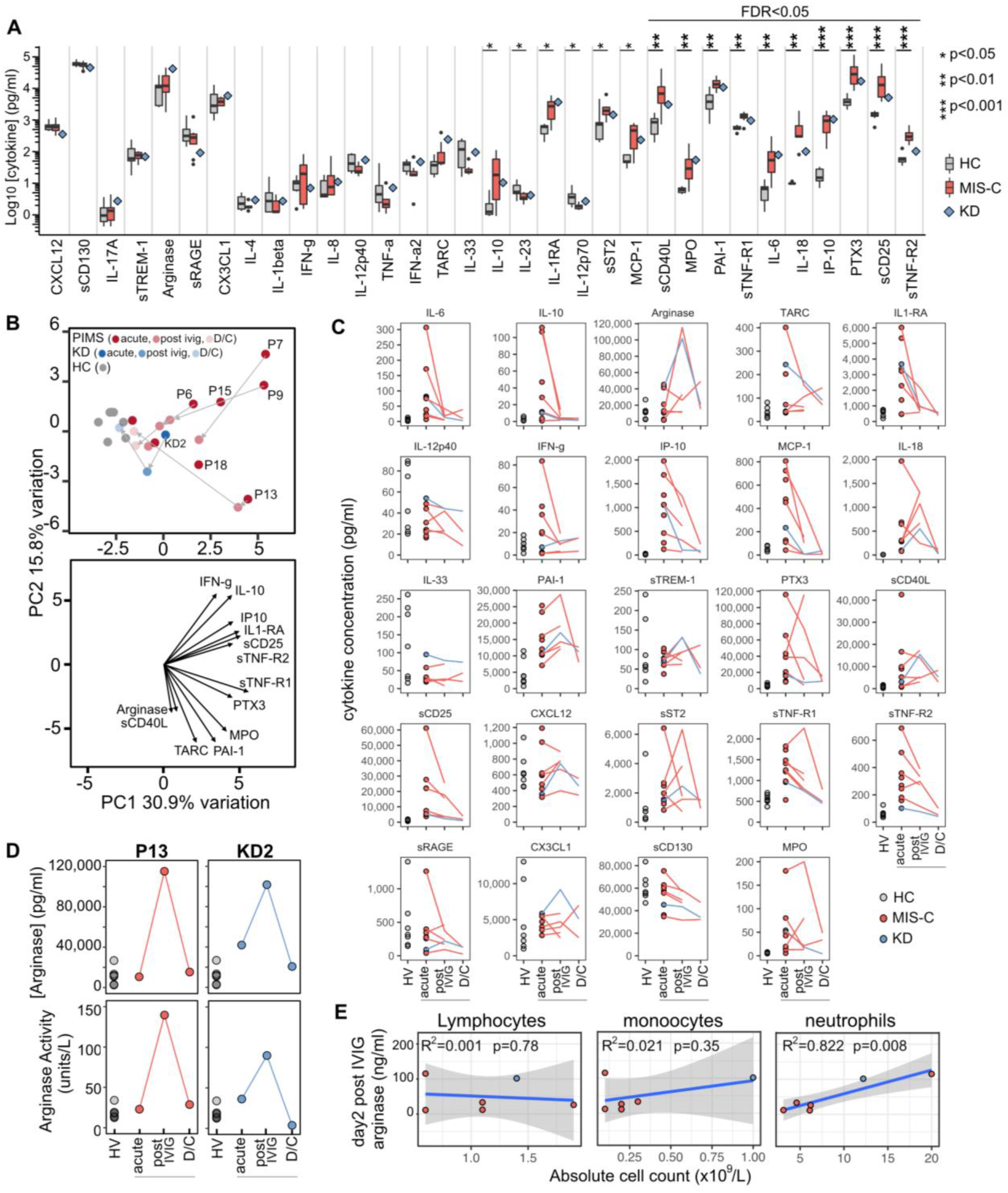
Analysis of cytokines in plasma samples from healthy children and patients. **A**. Levels of cytokines in plasma from seven healthy children or six MIS-C patients at the acute stage of disease are shown as box and whisker alongside a blue diamond indicating results from a single KD patient also at the acute stage. Results of Wilcoxon rank-sum tests comparing the frequency of each cluster in healthy children and MIS-C patients are indicated by: * p<0.05, ** p<0.01, *** p<0.001. Non-significant results are not shown and emboldened p value symbols indicate significant results after 5% false discovery rate correction using the Benjamini-Hochberg method. **B**. Upper panel: principal component analysis biplot of cytokines. Lower panel: loading plot showing the top 13 features contributing to principal components one and two. **C**. Trajectory of cytokines over time for the same healthy children and patients shown in panel A at the acute, post-IVIG and PICU-discharge timepoints. No statistical testing was performed. **D**. Plots showing the concentration (upper panel) and enzyme activity (lower panel) of arginase over time in plasma samples from seven healthy children, MIS-C patient P13 and KD patient KD2. **E**. Results of linear regression analysis of the acute disease stage absolute counts of lymphocytes, monocytes or neutrophils against the plasma arginase concentration after IVIG treatment. The R^2^ and statistical significance of each regression model is shown on the plot with the shaded area indicating the 95% confidence interval.

Principal component analysis divided patients into two broad groups (**Figure 7B**). Acute MIS-C samples were most distant from the healthy children and almost all patients migrated towards the healthy state after IVIG, the exception being P13, a patient with particularly severe disease. Examining each soluble mediator over the disease course (**Figure 7C**) showed many decreased following treatment with IVIG or IVIG combined with steroids (**SI Figure 7**). These included both proinflammatory (IL-6, IP-10, MCP-1) and anti-inflammatory (IL-10, IL-1Ra) molecules. A notable exception was arginase, levels of which in the acute phase of disease were not significantly higher than those in healthy children but increased dramatically for MIS-C patient P13 and patient KD2 in their post IVIG sample. Patient P13 received IVIG and then steroids before this sample but patient KD2 received IVIG alone. The increased quantity of arginase in these patients’ plasma was confirmed to be enzymatically active in an independent assay (**Figure 7D**). Finally, across all patients we noted a significant positive correlation between post-IVIG arginase levels and pre-treatment absolute number of neutrophils (r=0.91, R^2^=0.822, p=0.008), the main source of arginase in humans^33^ but no correlation with lymphocytes or monocytes (**Figure 7E**).

## Discussion

MIS-C is a newly described rare distinct acute paediatric inflammatory illness occurring several weeks after primary SARS-CoV-2 infection. There are currently few reports investigating the immune system in MIS-C and these predominantly focus on the adaptive immune system with the limitation of some examining samples only collected after immune modulation therapy or from patients lacking SARS-CoV-2 antibodies^13,14^. We performed a detailed analysis of samples collected from MIS-C patients with confirmed SARS-CoV2 serology taken before treatment and thereafter. The clinical features of our MIS-C cohort were consistent with other studies: for example older than 5 years old, high blood levels of inflammatory markers, ferritinaemia, neutrophilia, lymphopaenia and increased numbers of plasmablasts ^2,3,12,13,34^ Neutrophilia has previously been reported to correlate with IVIG resistance in KD^35^. For MIS-C, our data shows increased neutrophil count is positively correlated with cardiac dysfunction, inflammation, and disease severity. All of the MIS-C patients were deficient for vitamin D, which is linked to greater disease severity in KD^36,37^ and inflammation^38^. The clinical significance of this finding remains to be determined since in the UK deficiency is common in the black and ethnic minority groups from which 15/16 of our MIS-C patients were from^39^.

Our high dimensional immune analysis provided three key insights into the immunopathology of MIS-C. First, the acute phase is a hyper-inflammatory state with coincident activation of both the innate and adaptive arms of the immune system. Although the frequency of monocytes was not altered in the blood, all MIS-C patients had highly activated classical monocytes. Neutrophils were also highly activated and these were present at high frequency in the circulation. A large proportion lacked CD10 indicating immaturity. Classical monocytes and neutrophils both strongly expressed CD64, a canonical marker of activation for these cell types but which also serves as the high-affinity Fc receptor^9^. Thus these cells could potentially be activated by autoantibodies that are reported to be present in MIS-C patients^11,14^. Finally, consistent with a recent report we found a substantial proportion of memory CD8 T-cells, but only a small proportion of memory CD4 T-cells, were activated in MIS-C patients^34^.

Second, acute MIS-C patients have elevated levels of both pro- and anti-inflammatory cytokines in their plasma. All patients in our cohort had raised IL-6, consistent with their raised levels of CRP that is produced by the liver in response to IL-6. We did not detect raised levels of IL-17A, TNF-a, IFN-ϒ or IL1-beta, results that differ from some groups^13,14,16,40^ but agree with others^10,11,40^. These differences may reflect differences in patient cohorts or assay sensitivity. However, we identified significantly increased levels of multiple other cytokines including: the chemokines IP10 and MCP1, the pro-inflammatory cytokines PTX-3, IL-18 and the anti-inflammatory cytokines IL-10, sTNF-R1, sTNFR2 and IL-1RA. We also detected raised levels of MPO, consistent with the strong myeloid cell activation present in our MIS-C patients, and PAI-1, a hallmark of endothelial dysfunction that is raised in multiple inflammatory conditions including trauma and sepsis and that amplifies neutrophil-mediated inflammation via multiple mechanisms^29,41^.

Third, over the disease course there were substantial immune changes. Only two days after IVIG infusion the proportion of activated cluster 20 classical monocytes had decreased substantially with a concomitant increase in cluster 17 classical monocytes. These cells were, however, still phenotypically distinct to cluster 17 cells present in healthy children. They expressed intermediate CD64, and are therefore activated, but also now expressed CD163, a marker of anti-inflammatory monocytes. Soluble CD163 is a marker of monocyte activation produced by monocytes co-expressing CD163 and the inflammation-induced protease TACE, which cleaves CD163 from the cell surface^42^. Therefore, the presence of CD163 on monocytes following IVIG could represent monocytes transitioning to a less-activated state, no longer shedding CD163 due to downregulation of TACE, or the establishment of a new population of monocytes with anti-inflammatory activity^43^.Neutrophils were also altered two days after-IVIG, with levels of CD64 decreased from the peak pre-treatment level although still clearly raised above levels seen in healthy children. These decreases in innate immune cell activation shortly after IVIG treatment were accompanied by contemporaneous decreases in levels of proinflammatory and anti-inflammatory cytokines. A notable exception was the immunosuppressive enzyme arginase, with levels and enzymatic activity increasing substantially in MIS-C patient 13 and KD patient 2 after treatment. These patients had different diseases but shared two features in common: pronounced neutrophilia and IVIG in their treatment regimen. KD2 did not receive steroids or any other immunomodulatory agent besides IVIG. Although we cannot identify the source of arginase in these patients, neutrophils are known to exert suppressive function via arginase in different inflammatory situations and are therefore a prime candidate^44,45^. The increase of arginase shortly after IVIG administration could suggest a potential new mode of action for this widely used drug.

While MIS-C shares clinical features with other paediatric inflammatory conditions such as KD and MAS, the extent to which this reflect the same underpinning mechanisms is unclear. Our study allows a direct comparison to be made between MIS-C and KD to identify similarities and key differences. Although we recognise the patient number was low, the KD patients we analysed had features consistent with multiple studies performed on KD including: lymphopaenia, neutrophilia; neutrophil activation; increased intermediate and non-classical monocytes; high CD64 expression on monocytes and neutrophils; undetectable or low levels of IL-1beta, IFN-ϒ and TNF-α; raised levels of IL6, IL18, PAI-1, sCD25, sTNF-R, IP10, MPO, PTX3 and therefore serve as exemplars of the disease in our analyses^27,28,30–32,46^. MIS-C patients had all these features in common with KD, however, there were two notable differences. The first is that MIS-C patients possessed increased frequencies of DN1 B-cells, a subset unaltered in KD^47^. DN1 B-cells are increased during pathogenic and protective immune responses and may represent precursor B-cells undergoing differentiation into memory B-cells^21,48^. The second is that MIS-C patients lacked the expansion of intermediate and non-classical monocytes that is a hallmark of KD^32,49^. The function of these monocyte subsets, normally rare in the peripheral blood, is still being defined but they are generally considered to play a role in tissue repair^50^. Of particular relevance for KD vasculitis, the expression of CX3CR1 on NCM (^19^ and our scRNAseq data) allows them to closely interact with the vasculature^51^. Whether the lack of these cells in MIS-C is the reason why these patients present with severe shock, which is rare in KD, requires further investigation.

In summary, our data shows MIS-C is associated with contemporaneous activation of classical monocytes, neutrophils and CD8+ memory T-cells, increased numbers of double-negative B-cells and raised levels of pro-but also anti-inflammatory cytokines. Almost all of these features were present in our KD patient and have been previously reported in larger KD cohorts. We also observed important differences that we postulate may contribute to the different clinical presentation for MIS-C, notably the cardiac dysfunction, hypotension and life-threatening shock. Occasionally patients with KD present with cardiogenic shock (Kawasaki Disease Shock Syndrome -- KDSS) similar to the MIS-C clinical presentation^52–54^. While MIS-C has been suggested to be a distinct disease, our data suggest the immunopathology of MIS-C and KD are fundamentally similar. We therefore suggest that MIS-C resembles KDSS.

Therapy for MIS-C has empirically followed protocols used to treat KD, including IVIG and targeted therapies that inhibit IL-1beta (Anakinra), TNF-α (Infliximab) or IL-6 (Tocilizumab) ^2,3,12^. Our MIS-C patients possessed low levels of IL1-β and TNF-α but high levels of their natural antagonists, IL1-RA and sTNF-r. Additional therapeutic inhibition of these pathways may therefore have limited benefit and IL-6 inhibition may therefore be preferred. Although observational studies, such as ours, cannot definitively prove causality, the consistent and rapid immune changes that occurred in our MIS-C and KD patients shortly after IVIG (which in most cases was given as single agent) are striking, supporting the use of IVIG in MIS-C.

## Methods

### Study design, participants and ethical approval

All patient samples were obtained Birmingham Children’s Hospital as part of a Health Research Authority approved study (TrICICL) reviewed and approved by South of Birmingham Research Ethics Committee (REC: 17/WM/0453, IRAS: 233593). Samples from seven healthy children (aged 12 years) were obtained via the Coronavirus Immunological Analysis study approved by North West - Preston Research Ethics Committee (REC: 20/NW/0240, IRAS: 282164). This study was performed in accordance with the declaration of Helsinki and written informed consent was obtained from all participants or their legal guardians.

### Blood Samples and PBMC Isolation

Peripheral blood samples from paediatric patients presenting with suspected MIS-C were collected in EDTA and serum vacutainer tubes and stored at room temperature before processing. Whole blood was preserved using Cytodelics stabiliser and peripheral blood mononuclear cells (PBMCs) isolated and cryopreserved. Plasma collected during PBMC isolation and serum separated by centrifugation were stored frozen at −80°C.

### scRNA sequencing

Cryopreserved PBMCs were recovered and feature barcoded using DNA-conjugated Totalseq-B antibodies (Biolegend): anti-human TCRg/d, anti-human CD45RO and the TotalSeq-B Human TBNK cocktail specific for CD3, CD4, CD8, CD11c, CD16, CD19, CD45 and CD56. Single cell libraries were constructed using 10X Genomics Single Cell 3’ Reagents Kits v3 according to manufacturer’s instructions and sequenced on an Illumina NextSeq 500 platform. Details of the data analysis is provided in supplementary methods.

### Mass Cytometry

Antibodies were purchased pre-conjugated from Fluidigm or unconjugated from Biolegend and conjugated in house using Maxpar reagents (Fluidigm). Whole blood samples stored in Cytodelics were recovered according to manufacturer’s protocol. Details of antibodies and the staining protocol are provided in supplementary methods.

### Mass Cytometry data analysis

Bead normalised data were analysed using Cytobank software. Singlet cells from healthy children or MIS-C patients at different clinical stages were examined individually or as concatenated FCS files comprising an equal number of cells from each individual. Each FCS file was then downsampled to 16,000 cells (mononuclear cells) or 22,000 cells (granulocytes) for analysis. Gating strategies are provided in supplementary Figure 10. Dimensionality reduction and clustering were performed in Cytobank using the ViSNE implementation of tSNE and FlowSOM respectively.

### Cytokine quantification

Cytokines were quantified in plasma using the BioLegend LEGENDplex macrophage panel, human inflammatory panel 1 and 2 assays using a BD Fortessa cytometer and analysed using the LEGENDplex data analysis software suite. Myeloperoxidase was measured in plasma by ELISA (Fisher Scientific).

### Data analysis and statistical testing

Data analysis was performed in R using the lm, PCAtools, marixStats and matrixTests packages. We report the significance of each test. Where necessary, we also show whether statistical test results are significant after controlling the false discovery rate using the Benjamini-Hochberg procedure. The details of each test are provided in figure legends. The correlation matrix was produced in R using publicly available code downloaded from GitHub (Wherry research group, University of Pennsylvania). Graphs were prepared using ggplot2 in R v.4.0.3 or Prism v.8 (GraphPad Software)

## Data Availability

RNAseq and mass data will be available upon article acceptance.

## Acknowledgements

We thank the children and their families for consenting to join this research study and Birmingham Women’s and Children’s Hospital Charity for funding the single cell RNA sequencing analysis. No other external funding was received. GT is a member of the United Kingdom Coronavirus Immunology Consortium and healthy children were recruited via an ethical approval obtained via the consortium. ES is supported by a CRUK Birmingham Centre clinical PhD fellowship (C17422/A27438). BS is funded by a National Institute for Health Research (NIHR) Clinician Scientist fellowship. PGM is supported by the European Regional Development Fund (No. CZ.02.1.01/0.0/0.0/16_019/0000868). The views expressed are those of the author(s) and not necessarily those of the NHS, the NIHR or the Department of Health and Social Care. We thank Genomics Birmingham (University of Birmingham) for generating the single cell sequencing data. We would like to acknowledge staff at the Birmingham Women’s and Children’s NHS Foundation trust, including Helen Winmill, Sarah Fox, Carly Tooke, Samantha Owen, Natalie Read, Julie Menzies, Fiona Reynolds, Jim Gray, Mitul Patel, Phillip Hurley, Tristan Ramcharan, Kavitha Masilamani, Habib Ali, Sakeena Samar, Penny Davis, Kathryn Harrison, William Coles, Deevena Chinthala, Heather Duncan, Nick Richens and Sanket Sontakke.

## Author Contributions

ES, PK and GT designed the research study. ES, NK, SF, AR and GT conducted the experiments and provided the reagents. ES, AR, EAA, AC, PD, SH, DJ, HKK, SM, PN, BRS, SW, GT helped with data collection. ES, EF, PV, RS, SO and GT analysed data. All authors contributed to writing the manuscript.

## Supplementary Information

### Overlap with prior publications

Initial presenting features during PICU of five patients with MIS-C have been reported in a recent study^S1^. Cardiac features in five patients and antibody profiles of four patients have also been presented in our single centre reports^S2,S3^. The single cell RNA sequencing, mass cytometry and cytokine analyses reported in the current manuscript have not been published previously.

### Community SARS-CoV-2 data

Time series of positive cases by testing date for England was downloaded from the UK Government Department of Social Care and Public Health England website: https://www.gov.uk/guidance/coronavirus-covid-19-information-for-the-public#time-series-documents. Data shown are results for area ‘Birmingham’.

### Blood Samples and PBMC Isolation

Peripheral blood samples from paediatric patients presenting with suspected MIS-C were collected in EDTA tubes and stored at room temperature before processing. Peripheral blood mononuclear cells (PBMCs) were isolated using SepMate tubes (Stemcell Technologies) as per the manufacturer’s protocol. PBMCs were cryopreserved in medium containing DMSO ^S4^ before transfer to liquid nitrogen for long-term storage. All blood samples used for scRNAseq analysis were processed within 4 hours of phlebotomy.

### Clinical laboratory and antibody data generation

Clinical laboratory tests were performed at the Birmingham Children’s Hospital’s clinical laboratory. Lymphocyte subset enumeration was performed by the University of Birmingham Clinical Immunology Service using BD Trucount™ tubes (CD3,4,8,19,15,56) and analysed using a BD FACS Canto II cytometer. Paediatric reference values were obtained from previously published data ^S5^. Screening of patients for anti-SARS-CoV-2 spike protein responses was performed as described ^S3^. Further analysis of anti-SARS-CoV2 IgM, IgA and IgG subclasses was performed using ELISA plates coated with stabilized, trimeric spike glycoprotein truncated at the transmembrane region (The Binding Site, UK). Serum was pre-diluted at a 1:40 dilution using a Dynex Revelation automated liquid handler (Dynex, USA). Antibodies were detected using sheep-anti-human HRP-conjugated polyclonal antibodies against IgG (1:16,000), IgA (1:2000), and IgM (1:8000), TMB core and orthophosphoric acid as a stop solution (all from The Binding Site, UK). Optical densities at 450nm were measured using the Dynex Revelation automated liquid handler. IgG, IgA, and IgM ratio-cutoffs were determined based on running 90 pre-2019 negative serum samples. Ratio values > 1, are classed as positive and ratio values < 1 are classed as negative for anti-SARS-CoV-2 spike IgG, IgA or IgM antibodies.

### ScRNAseq data generation

For scRNAseq analysis cryopreserved PBMCs were recovered and feature barcoded using the following DNA-conjugated Totalseq-B antibodies (Biolegend): anti-human TCRg/d, anti-human CD45RO and the TotalSeq-B Human TBNK cocktail designed to react against CD3, CD4, CD8, CD11c, CD16, CD19, CD45 and CD56. Single cell libraries were constructed using 10X Genomics Single Cell 3’ Reagents Kits v3 according to manufacturer’s instructions and sequenced on an Illumina NextSeq 500 platform.

### ScRNAseq Data Pre-processing and Quality Control

Processing of raw reads including 10X barcode-aware demultiplexing from BCL to FASTQ files, transcriptome alignment to human genome assembly GRCh38 and unique molecular identifier (UMI) counting were performed using the 10X Cell Ranger pipeline version 3.1.0 with GRCh38-version 3.0.0 as the reference. Seurat v3.1.5 ^S6^ was used for sample merging, quality control (QC), clustering and reporting. Both gene expression (RNA) and antibody-derived tag (ADT) assays were loaded from the CellRanger count platform into a Seurat object for each sample excluding cells with less than 200 genes and features detected in less than 3 cells. The four samples were then merged to create a single aggregated object. QC was conducted on both assays separately. For the RNA assay, to mitigate the influence of sex on clustering and differential expression, the XIST and RSP4Y1 genes were removed. Cells with a number of features between 200 and 600, counts of greater than 1000 and mitochondrial percentage less than 15% were kept for further processing. The gene expression data was normalised using the ‘LogNormalize’ method with the scale.factor set to the default 10000. 1500 variable features were identified using the ‘vst’ method of the ‘FindVariableFeatures’ function. The assay was finally scaled with the number of counts and mitochondrial percentage variables regressed. The ADT assay was first used to identify dead cells or cell doublets by removing double positive instances of lineage markers according to the following criteria: CD14 > 60 & CD19 > 50, CD19 > 40 & CD3 > 40, CD14 > 80 & CD3 > 50 and CD3 < 50 & gamma-delta > 400. The assay was then centred log-ratio (CLR) normalised and scaled with default parameters. The remaining object comprised 10,031 cells with 17,583 features.

### PBMC Clustering of scRNAseq data

Both the RNA and ADT assays were used to compute 50 principal components. These principal components were then used as input into the ‘FindNeighbours’ function and subsequently the ‘FindClusters’ function with resolution set to the default value of 1, resulting in 19 clusters which were visualised using tSNE. The clusters were manually annotated from expression of lineage markers both at the transcript (RNA) and protein (ADT) levels. To check our samples for potential T-cell activation we evaluated our data for expression of genes previously identified by scRNAseq analysis as being upregulated upon T-cell stimulation ^S7^.

### Monocyte Clustering of scRNAseq data

To examine the monocyte populations in finer detail, we extracted clusters 4, 6, 9, 10 and 13 from Figure 1E using the ‘Subset’ function. As monocytes were abundant within each sample, all monocyte clusters were included (2248 monocytes extracted). Both assays were normalized separately (RNA: ‘LogNormalize’ & ADT: ‘CLR’). The ‘vst’ method was used to find the 2000 most variable features within the RNA assay and the data was scaled with the variables ‘number of counts’ and ‘percent mitochondria’ regressed. The ADT assay was scaled with default parameters. In this instance, 100 principal components were used in the ‘RunPCA’ function with the top 50 of those components used to find cell neighbours and clusters. The UMAP reduction was selected here as the clusters visually displayed a better path from classical to intermediate to non-classical monocytes over the tSNE reduction. Nine clusters were originally generated with further assessment resulting in the identification of seven monocyte clusters. CD14 and CD16 markers from the ADT assay were used to classify classical (CD14+), non-classical (CD16+) and intermediate (CD14+CD16+) monocyte populations. Further differential expression analysis of the RNA assay was used to define cellular function within each cluster.

### Gene Ontology

Differential gene expression was conducted between the acute samples and the convalescent sample for CD14+ monocytes, NK, CD8+ T cells, CD4+ T cells and B cells with the ‘FindMarkers’ function in Seurat using MAST ^S8^. Both up and down regulated genes were identified. Gene ontology was conducted with the resulting gene lists using EnrichR ^S9^, focusing on the biological processes subset (‘GO_Biological_Processes’). Go terms were simplified using Revigo ^S10^

### Mass Cytometry staining protocol

Whole blood samples stored in Cydotelics stabiliser media were thawed and processed as per manufacturer’s protocol. For staining, a master-mix of all 38 phenotyping antibodies was prepared by adding the appropriate pre-tested dilutions into filtered cell stain media (CSM - phosphate buffered saline + 0.5% Foetal Bovine Serum + 0.02% sodium azide). After incubating with FC block (Biolegend) for 10 minutes, antibodies were added and incubated for further 30 minutes. The samples were washed twice with CSM buffer then fixed overnight with freshly prepared 1.6% formaldehyde. The following day, cells were incubated with iridium intercalator solution (Fluidigm) for one hour then analysed on a Fluidigm Helios mass cytometer using an acquisition rate below 500 events per second. Immediately prior to acquisition cells were washed once in CSM buffer and twice in deionised water. Prior to acquisition, each sample was reconstituted in deionised water spiked with Four Element Calibration Beads (Fluidigm) and filtered through a 70μm cell strainer. Data was acquired on a Helios mass cytometer (Fluidigm) using an acquisition rate below 500 events per second. Resultant FCS data-files were then normalised using the CyTOF data acquisition software (Fluidigm). Normalised data were uploaded to Cytobank for further analysis including doublet discrimination and analysis by manual gating and unsupervised methods.

### Principal Component Analysis

Principal component analysis was performed using the PCAtools package in R. For the PCA analysis of clinical laboratory data presented in Figure 1F, although data from individual healthy donors was unavailable each feature had a well-defined normal range which varied depending on the age and sex of the child. Therefore, we generated synthetic healthy donor controls as follows. For each MIS-C or KD patient we generated ten synthetic healthy control donors by randomly selecting values from the normal range defined for that patient’s age and sex for each of the different features. Plotting each synthetic healthy donor with transparency produced a density map of normality for the entire patient cohort (shown in grey), against which the patients could be compared (Figure 1F). For cytokines (Figure 7) the PCA plot was prepared using the 24 features with highest variance (variance > 300).

