## Supplementary material for "The innate and adaptive immune landscape of SARS-CoV-2-associated Multisystem Inflammatory Syndrome in Children (MIS-C) from acute disease to recovery"

**Supplementary Figures and Tables**

**SI Table 1**. Demographic and clinical data for the 16 patients with MIS-C and 2 patients with Kawasaki disease (KD) recruited to the study. KD patients are indicated by grey background. PCR: polymerase chain reaction. D=Diarrhoea, V=Vomiting, Neg= Negative, Pos= Positive, M=Male, F=Female. Cardiac involvement includes abnormal ECG and/or echocardiography findings.

| ID | Age (years) | Sex | Ethnicity | Fever | Rash | Lymph-adenopathy | Conjunctivitis non-exudated | Mucosal changes | Peripheral changes | GI features | Cardiac involvement | PCR for SARS-CoV-2 | Serology for SARS-CoV-2 | Diagnosis |
|---|---|---|---|---|---|---|---|---|---|---|---|---|---|---|
| 1 | 0-4 | M | Asian British | Yes | Yes | No | No | No | No | No | NO | Neg x3 | Not done | KD |
| 2 | 0-4 | M | White British | Yes | Yes | Yes | Yes | Yes | Yes | Yes, V | NO | Neg x1 | Negative | KD |
| 3 | 5-10 | M | Black British-African | Yes | No | No | No | No | No | Yes, V | YES | Neg x3 | Positive | MIS-C |
| 4 | 5-10 | M | Asian British-Pakistani | Yes | Yes | No | Yes | Yes | Yes | Yes, abdominal pain, V | NO | Neg x1 | Positive | MIS-C |
| 5 | 5-10 | M | Asian British-Pakistani | Yes | Yes | Yes | Yes | Yes | No | Yes, acute abdomen, pain, V | Yes | Neg x3 | Positive | MIS-C |
| 6 | 5-10 | F | Black British | Yes | Yes | No | Yes | Yes | No | Yes, amdominal pain, D | Yes | Pos x1 Neg x2 | Positive | MIS-C |
| 7 | 5-10 | F | Asian British-Indian | Yes | Yes | No | No | No | No | Yes, abdominal pain, V | NO | Pos x2 | Positive | MIS-C |
| 8 | 5-10 | M | Asian British-Indian | Yes | No | No | No | No | No | Yes, acute abdomen, pain, D&V | YES | Neg x3 | Positive | MIS-C |
| 9 | 5-10 | F | Asian British-Pakistani | Yes | Yes | No | No | No | No | Yes, pain, D&V | YES | Negx2 | Positive | MIS-C |
| 10 | 5-10 | F | Black British-African | Yes | Yes | No | Yes | Yes | Yes | Yes,D&V | YES | Neg x2 | Positive | MIS-C |
| 11 | 5-10 | M | Black British-Carribean | Yes | Yes | No | Yes | Yes | Yes | Yes, abdominal pain, D&V | YES | Neg x2 | Positive | MIS-C |
| 12 | 5-10 | M | Asian British-Baghladeshi | Yes | No | No | Yes | No | Yes | Yes, abdominal pain, D&V | YES | Neg x2 | Positive | MIS-C |
| 13 | 5-10 | F | Black British-African | Yes | Yes | Yes | No | Yes | No | Yes, acute abdomen, D&V | YES | Neg x2 | Positive | MIS-C |
| 14 | 11-15 | F | Mixed White Black-Caribbean | Yes | No | No | No | No | No | Yes, admominal pain, D | YES | Neg x3 | Positive | MIS-C |
| 15 | 11-15 | M | Mixed White Black- Afro Caribbean | Yes | No | No | No | No | No | Yes, abdominal pain, D&V | YES | Neg x2 | Positive | MIS-C |
| 16 | 11-15 | F | White-Romanian | Yes | Yes | Yes | Yes | No | No | Yes, abdominal pain, V | YES | Neg x2 | Positive | MIS-C |
| 17 | 11-15 | F | Black British-Caribbean | Yes | Yes | No | No | No | No | Yes, D&V | YES | Neg x3 | Positive | MIS-C |
| 18 | 11-15 | M | Asian British-[illegible] | Yes | Yes | No | Yes | No | No | Yes, [illegible] | NO | Neg x3 | Positive | MIS-C |

**RCPCH 2020 guidelines for PIMS-TS**: A child presenting with persistent fever, inflammation (neutrophilia, elevated CRP and lymphopenia) and evidence of single or multi-organ dysfunction (shock, cardiac, respiratory, renal, gastrointestinal or neurological disorder). This may include children meeting full or partial criteria for Kawasaki disease. Exclusion of any other microbial cause, including bacterial sepsis, staphylococcal or streptococcal shock syndromes, infections associated with myocarditis such as enterovirus (waiting for results of these investigations should not delay seeking expert advice). SARS-CoV-2 PCR testing may be positive or negative

**SI Table 2**. Antibody Markers used for Mass cytometry

| Metal | Marker | Clone | Source |
|---|---|---|---|
| 89Y | CD41/42a61 | A2A9/6 & REA209 | Biolegend & Miltenyi |
| 106Cd | CD16 | 3G8 | Biolegend |
| 110Cd | CD14 | RMO52 | Beckman Coulter |
| 113Cd | CD2 | TS1/8 | Biolegend |
| 114Cd | CD8 | SK1 | Biolegend |
| 115In | CD57 | HCD57 | Biolegend |
| 116Cd | CD36 | 5-271 | Biolegend |
| 139La | FCeR1 | AER-37 | Biolegend |
| 141Pr | CD45 | H130 | Fluidigm |
| 142Nd | CD19 | HIB19 | Fluidigm |
| 144Nd | CD32 | FUN-2 | Biolegend |
| 145Nd | CD4 | RPA-T4 | Fluidigm |
| 146Nd | IgD | IA6-2 | Fluidigm |
| 147Sm | CD11c | 5-HCL-3 | Biolegend |
| 148Nd | CD69 | REA824 | Miltenyi |
| 149Sm | CD64 | 10.1 | Biolegend |
| 150Nd | CD62L | DREG56 | Biolegend |
| 151Eu | CD123 | 6H6 | Biolegend |
| 155Gd | CD45RA | HI100 | Fluidigm |
| 156Gd | CD177 | MEM-166 | Biolegend |
| 159Tb | CD86 | IT2.2 | Biolegend |
| 160Gd | CD39 | A1 | Fluidigm |
| 161Dy | CD163 | GHI/61 | Biolegend |
| 162Dy | CD55 | IS11 | Biolegend |
| 163Dy | CD56 | NCAM16.2 | Fluidigm |
| 164Dy | CD95 | DX2 | Biolegend |
| 166Er | CD35 | E11 | Biolegend |
| 167Er | CD27 | L128 | Fluidigm |
| 168Er | CD10 | H10a | Biolegend |
| 169Tm | CD25 | 2A3 | Fluidigm |
| 173Yb | CD3 | UCHT1 | Biolegend |
| 174Yb | CD40 | HB14 | Biolegend |
| 175Lu | CXCR4 | 12G5 | Fluidigm |
| 176Yb | CD63 | H5C6 | Biolegend |
| 194Pt | CD66b | 6/40c | Biolegend |
| 195Pt | CD235 | HI264 | Biolegend |
| 196Pt | CD38 | HIT2 | Biolegend |
| 198Pt | HLA-DR | L243 | Biolegend |

**SI Figure 1:** Monocytes, Lymphocytes, Granulocytes absolute counts and CRP is shown for each individual patient over the course of time in the hospital. Treatment is indicated as follows: ivig= Intravenous Immunoglobulin, S= IV Methylprednisolone and Toc=Tocilizumab.

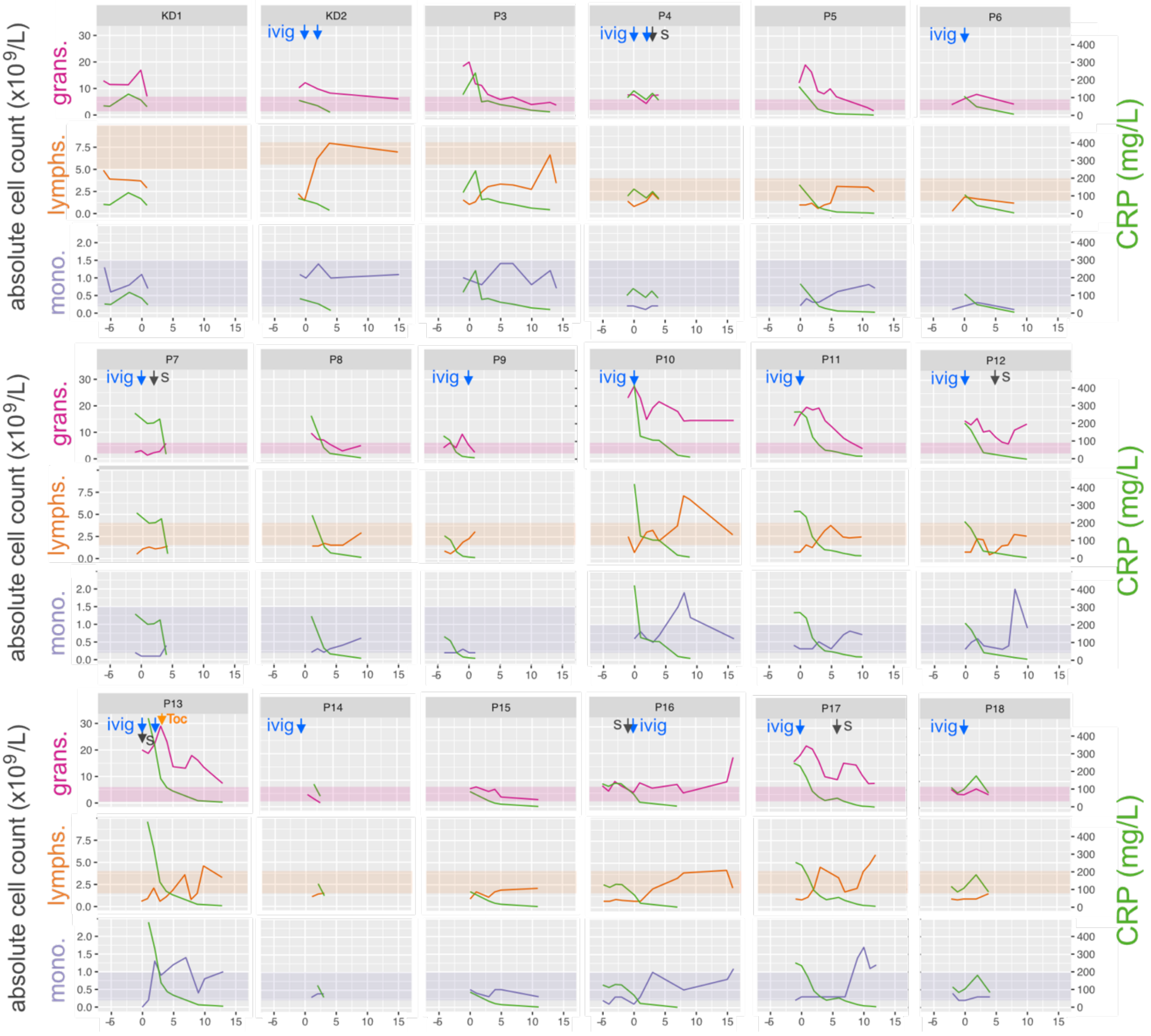

**SI Figure 2.** Gene expression profile of 19 clusters generated from patient PBMC samples.

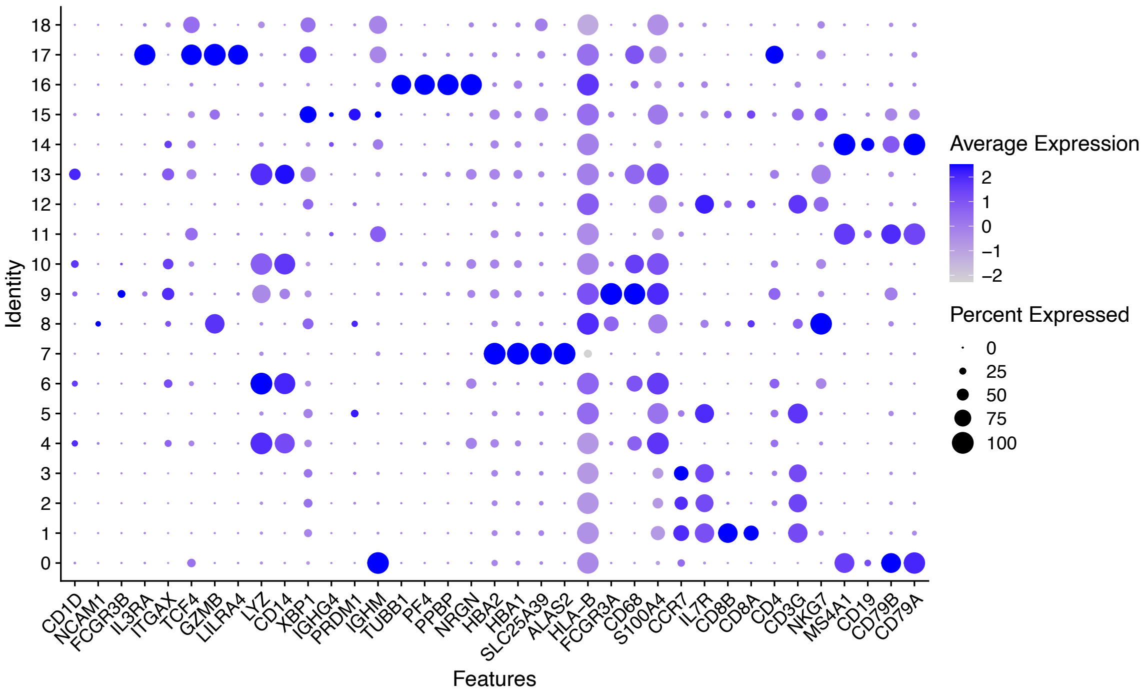

**SI Figure 3.** Heatmap showing scaled expression of discriminative genes sets for each PBMC cluster. Only the top five discriminative genes per cluster are shown.

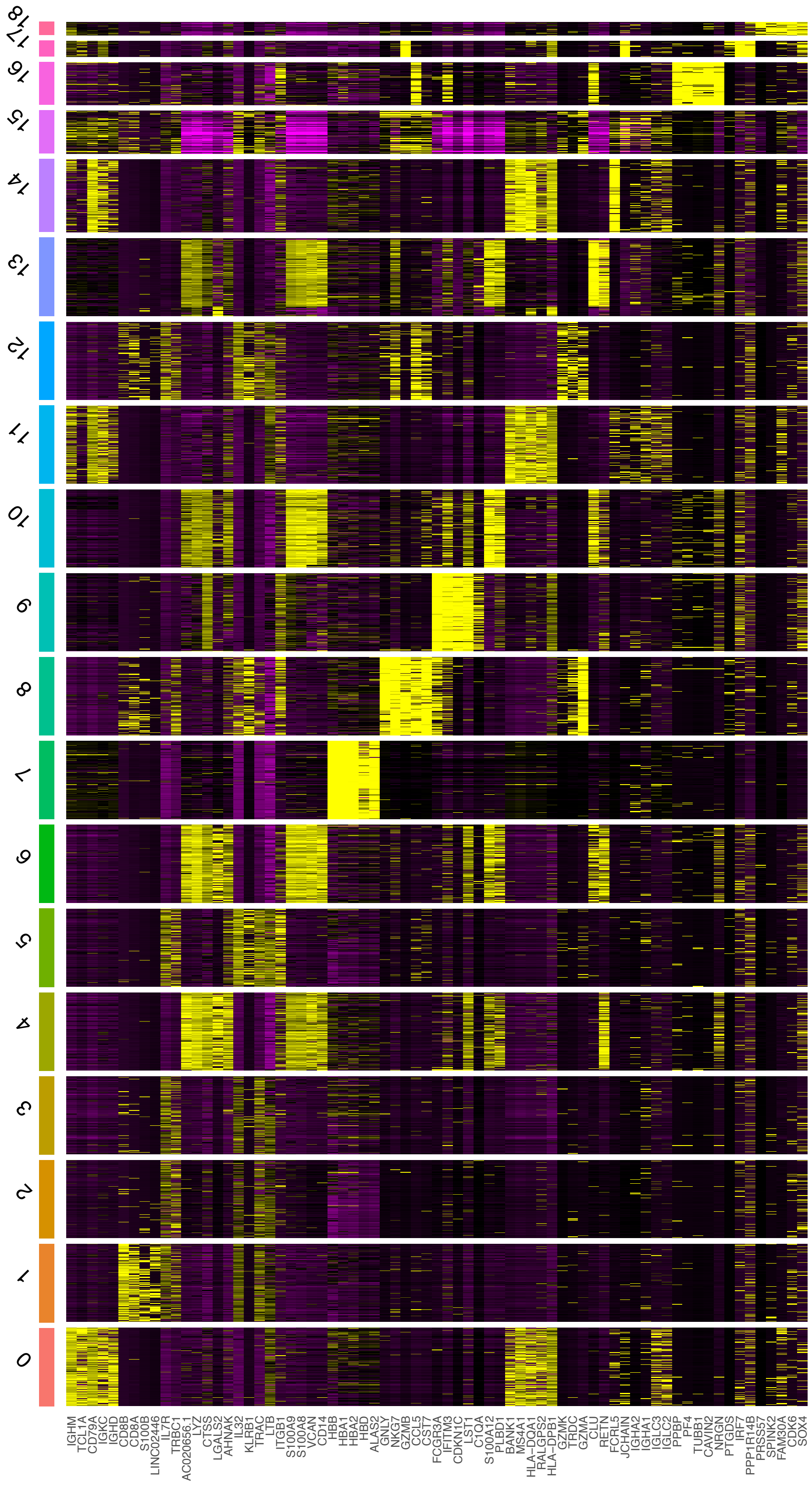

**SI Figure 4**. RNA expression and protein levels in patient PBMCs.
tSNE representation of combined PBMCs from all four patient samples coloured by the level of RNA per cell or level of protein per cell measured using antibody derived tags.

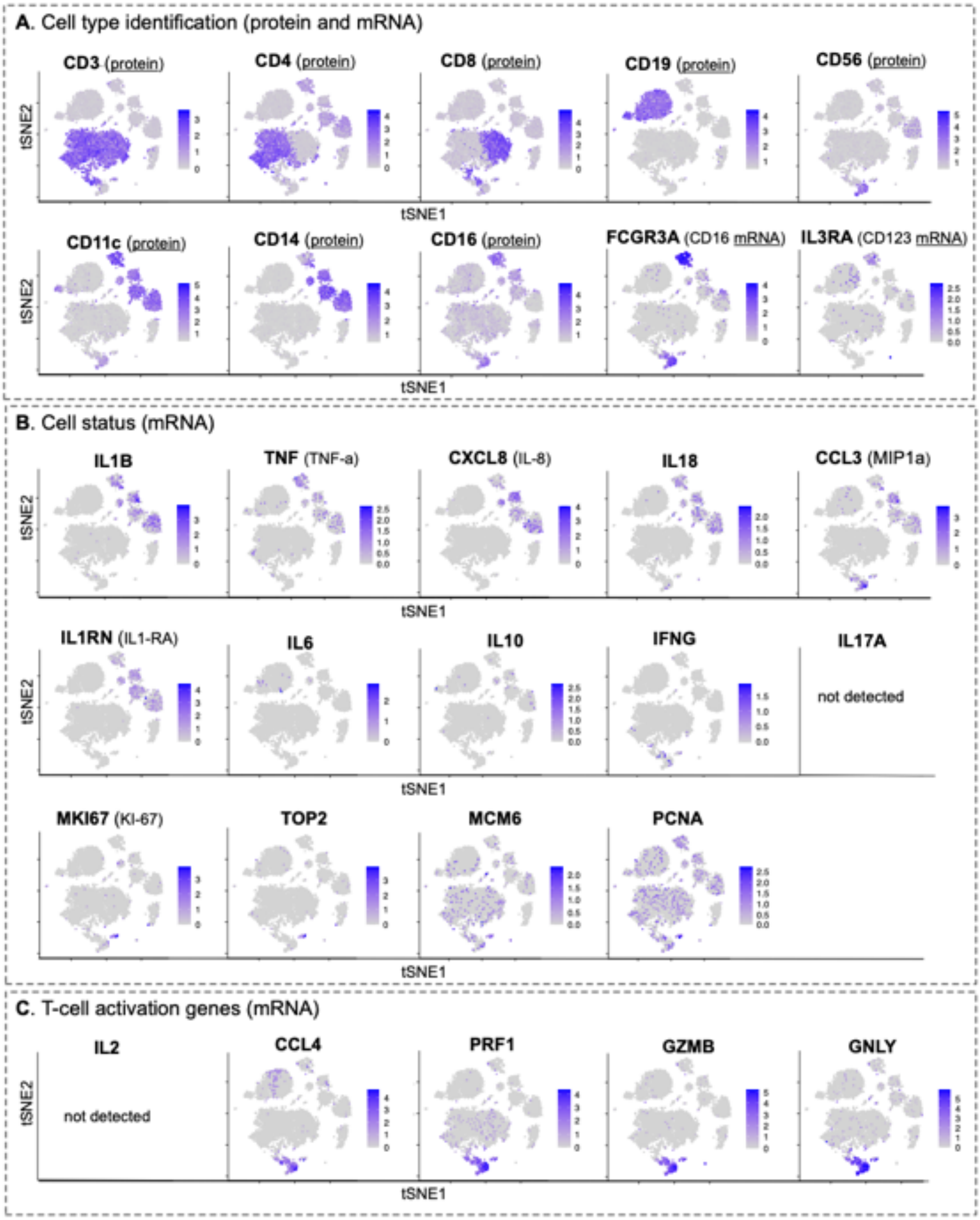

**SI Figure 5.** Heatmap showing scaled expression of discriminative genes sets for each of the new clusters generated by reclustering of cells from the originally identified monocyte clusters. Only the top five discriminative genes per cluster are shown.

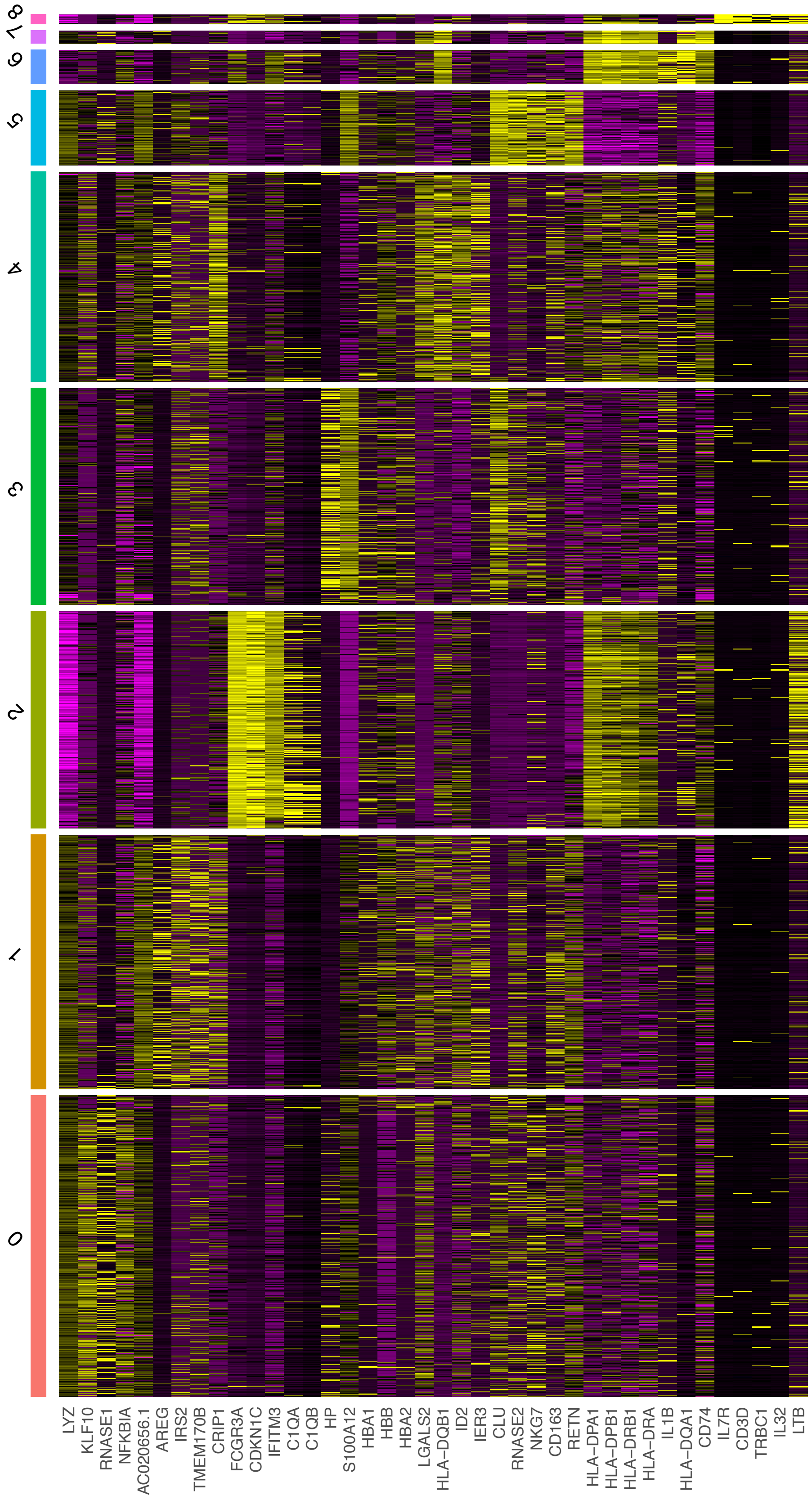

**SI Figure 6.** RNA expression and protein levels in patient monocytes. UMAP representation of monocytes from all four patient samples coloured by the level of RNA per cell or level of protein per cell measured using antibody derived tags.

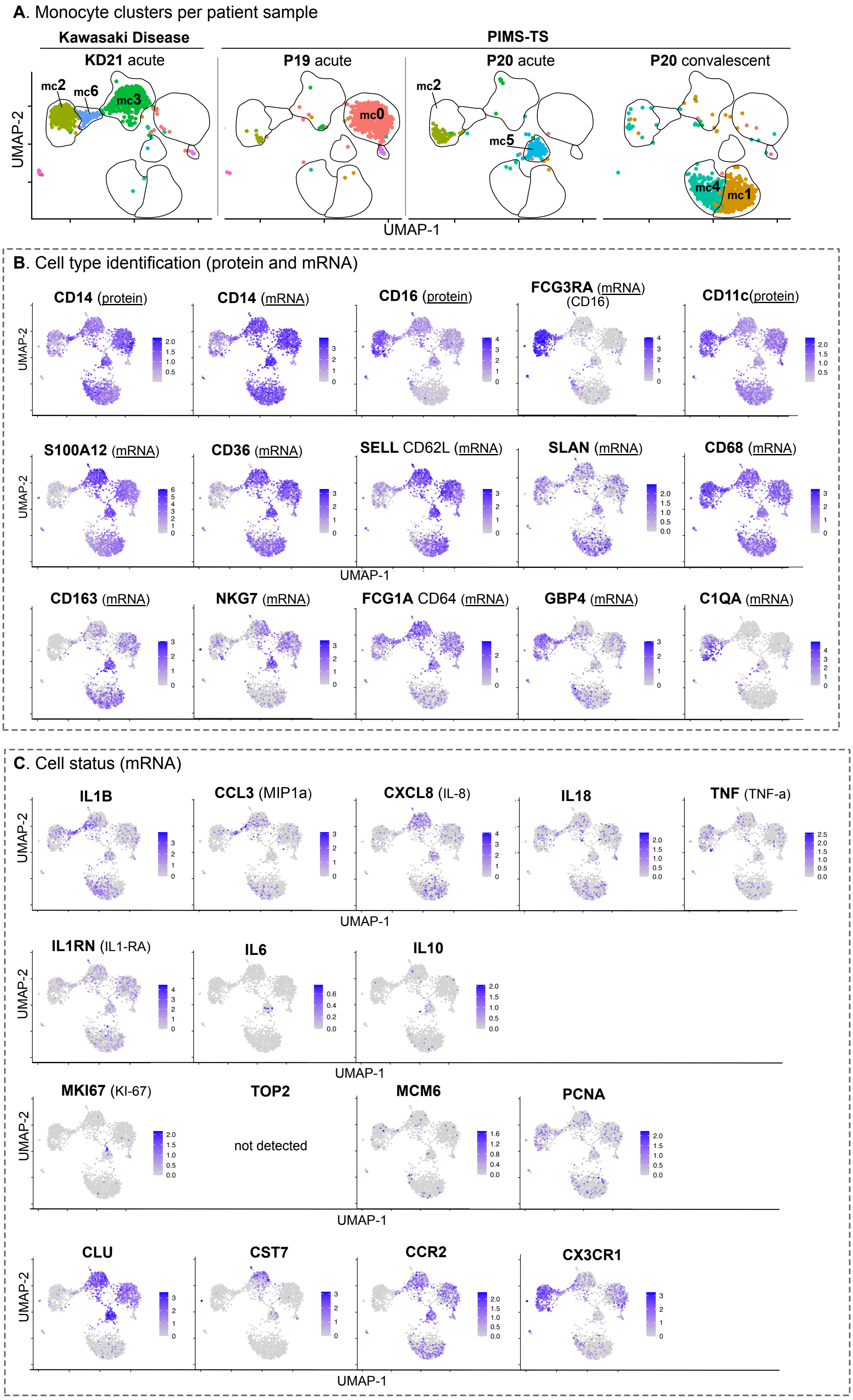

**SI figure 7.** Key events for each patient. Length of stay in hospital, treatments received and sample collection timings are shown.

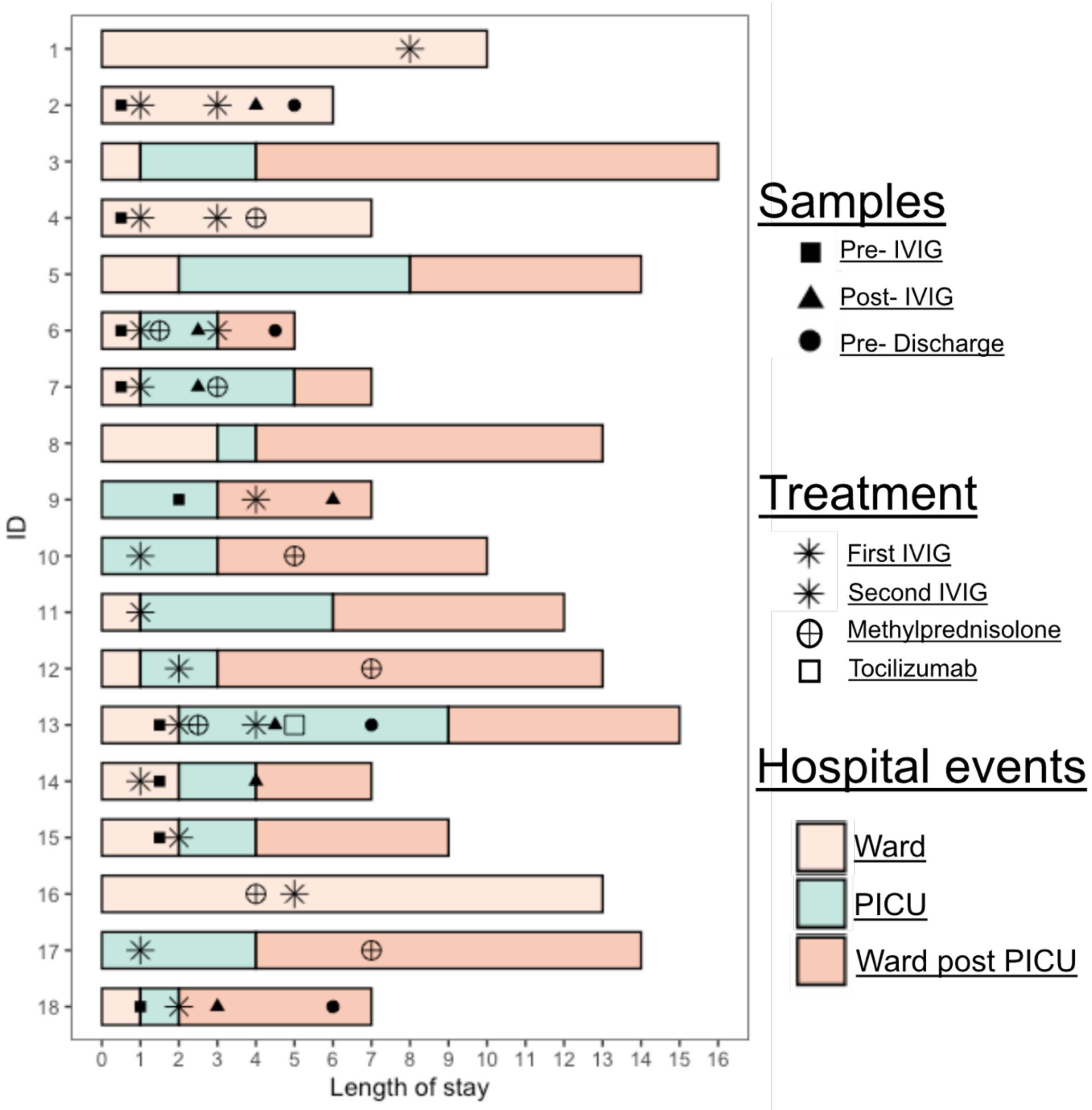

**SI Figure 8.** Heatmap showing the median mass intensity of each marker in each of the mononuclear cell clusters. The data shown in the figure was produced from six acute stage MIS-C patients (data from each patient combined together to produce the figure).

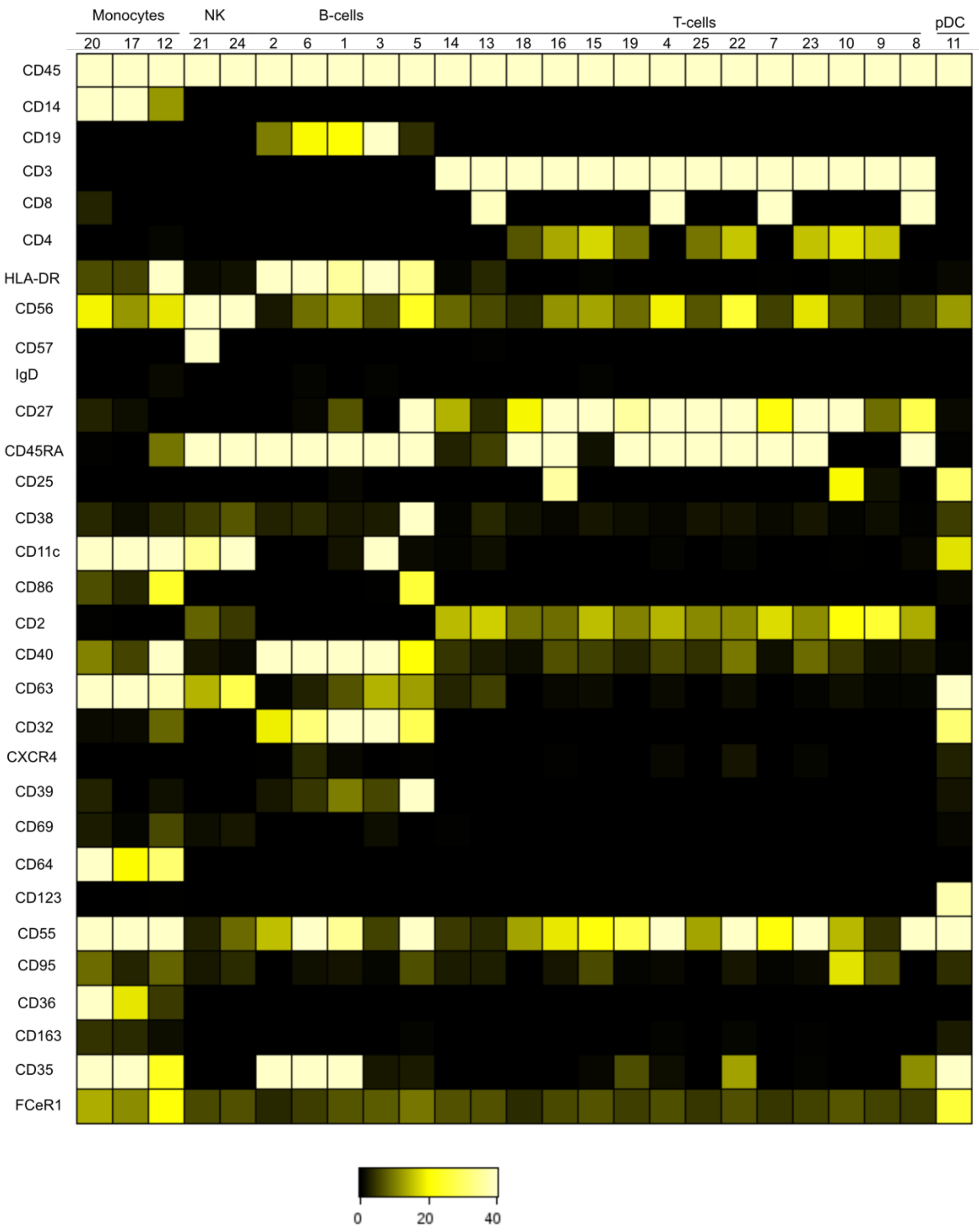

**SI Figure 9:** Biaxial plots of CD163 and CD64 expression for each individual for metacluster 17 (n=7 healthy donors, 6 MIS-C patients). D/C indicates sample taken at discharge (SI Figure 7).

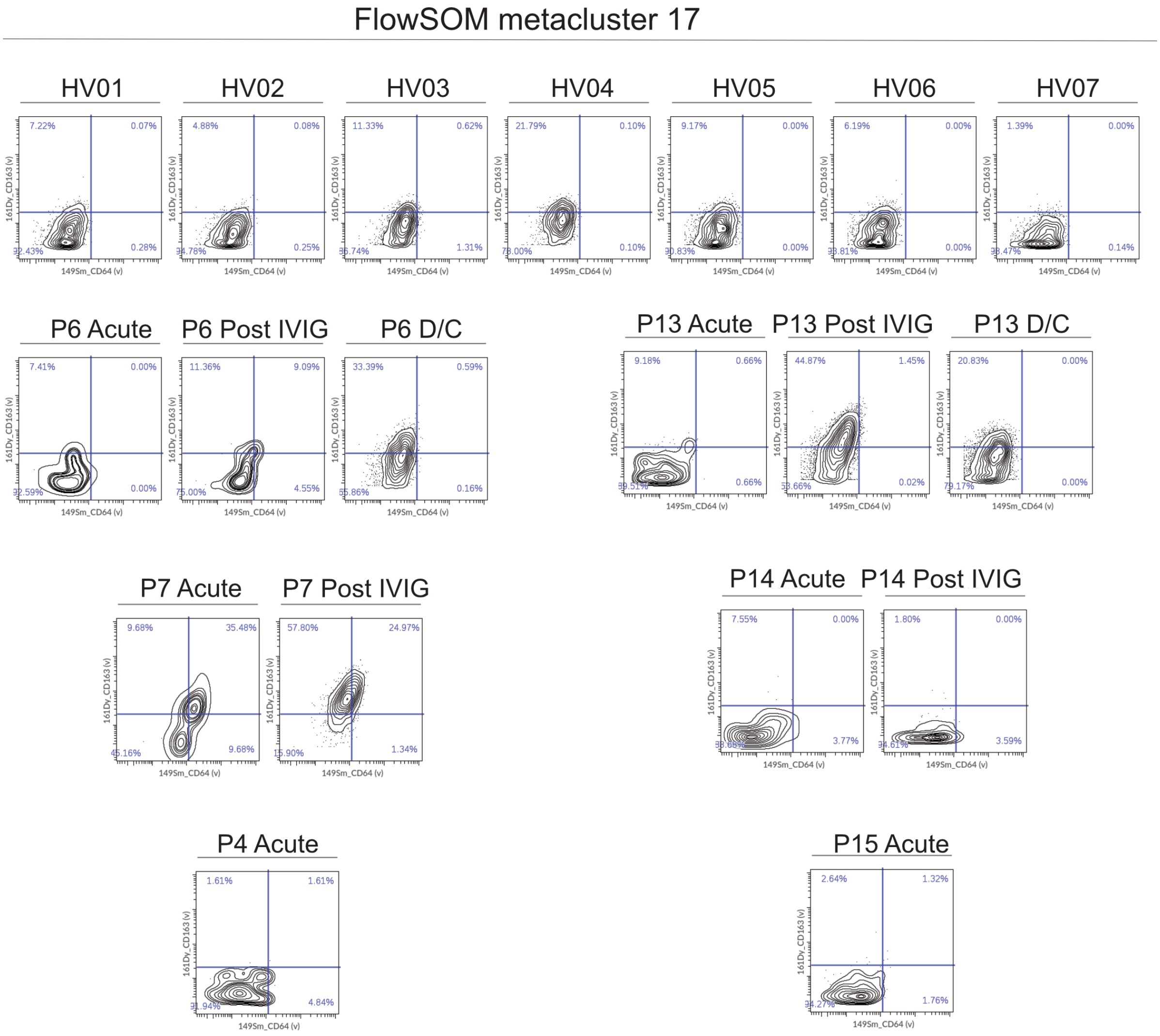

**SI Figure 10**. **A.** Gating strategy for major B and T cells subsets. **B.** Gating strategy for HLA-DR positive cells in the 4 subsets of CD8+ and CD4+ T-cells. **C.** Frequency of CD4+ and CD8+ T-cells within each of the four subsets for each individual healthy control and MIS-C acute patient. Statistical significance was determined by Wilcoxon test between healthy children and MIS-C patients at the acute stage of disease. ns: no significant difference.

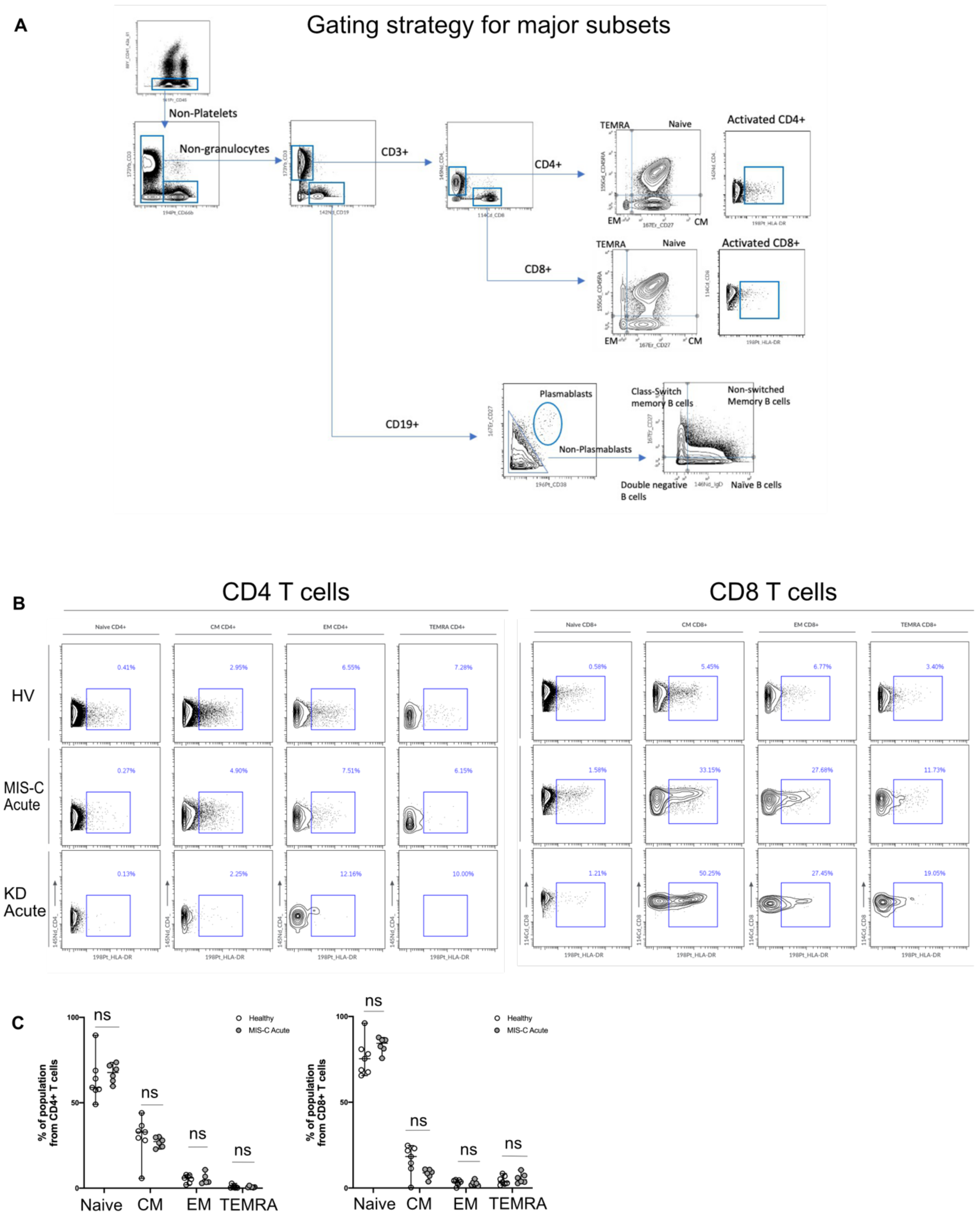
